# The effect of maternal BMI, smoking and alcohol on congenital heart diseases: a Mendelian randomization study

**DOI:** 10.1101/2022.01.27.22269962

**Authors:** Kurt Taylor, Robyn E. Wootton, Qian Yang, Sam Oddie, John Wright, Tiffany C Yang, Maria Magnus, Ole A. Andreassen, Maria Carolina Borges, Massimo Caputo, Deborah A Lawlor

## Abstract

**Background:** Congenital heart diseases (CHDs) remain a significant cause of infant morbidity and mortality. Epidemiological studies have explored maternal risk factors for offspring CHDs, but few have used genetic epidemiology methods to improve causal inference.

**Methods:** Three birth cohorts, including 38,662 mother/offspring pairs (N = 319 CHD cases) were included. We used Mendelian randomization (MR) analyses to explore the effects of genetically predicted maternal body mass index (BMI), smoking and alcohol on offspring CHDs. We generated genetic risk scores (GRS) using summary data from large scale genome-wide association studies and validated the strength of the genetic instrument for exposure levels during pregnancy. Logistic regression was used to estimate the odds ratio (OR) of CHD per 1 standard deviation (SD) change in GRS. Results for the three cohorts were combined using random-effects meta-analyses. We performed several sensitivity analyses including multivariable MR to check the robustness of our findings.

**Results:** The GRSs associated with the exposures during pregnancy in all three cohorts. The associations of the GRS for maternal BMI with offspring CHD (pooled OR (95% confidence interval) per 1SD higher GRS: 1.01 (0.90, 1.13)) and lifetime smoking (pooled OR: 0.97 (0.87, 1.08)) were close to the null, though with wide confidence intervals. We observed weak evidence of an increased odds of offspring CHDs with increase in the maternal GRS for alcoholic drinks per week (pooled OR: 1.09 (0.98, 1.22)). Sensitivity analyses yielded similar results.

**Conclusions:** Our results do not provide robust evidence of an effect of maternal BMI or smoking on offspring CHDs. However, results were imprecise. Our findings, including the potential effect of maternal alcohol intake on offspring CHD need to be replicated, and highlight the need for more and larger studies with maternal and offspring genotype and offspring CHD data.

## Introduction

Congenital heart diseases (CHDs) are the most common congenital anomaly, affecting 6-8 per 1000 live births and 10% of stillbirths ^1^. CHDs are a leading cause of childhood mortality and many CHD patients experience health problems that persist into adulthood ^2,3^. The causes of CHDs are largely unknown, but the pregnancy environment (intrauterine factors) may play a role in the underlying pathophysiology ^4^. Identifying modifiable risk factors for CHDs is important for improving aetiological understanding and developing preventive interventions to reduce disease burden.

Several modifiable maternal characteristics have been found to be associated with increased risk of CHDs, including maternal pre/early pregnancy body mass index (BMI) ^5–7^, smoking ^8^ and alcohol ^9^ consumption in pregnancy. The causal relevance of the results from meta-analyses is unclear, due to many studies not controlling for key confounders and for the risk of residual confounding. Previously, using parental negative exposure control analyses, we found that positive associations between maternal overweight and obesity with offspring CHDs may be being driven by confounding factors ^10^. This work found some evidence of an intrauterine effect of maternal smoking on offspring CHDs. For alcohol consumption, results were inconclusive due to limited data ^10^. Negative control analyses attempt to address the issue of residual confounding in observational studies ^10,11^, but rely on assumptions that cannot be empirically verified, such as it being implausible that the exposure in the father (e.g. their smoking) could influence offspring CHD risk to a similar magnitude of any effect in mothers.

Mendelian randomization (MR) uses genetic variants as instrumental variables (IVs) to test causal effects in observational data ^12^. The key assumptions for MR are: (i) relevance assumption - the genetic instruments are robustly associated with the exposure, (ii) independence assumption - there is no confounding of the genetic instrument-outcome association, (iii) exclusion restriction criteria - the genetic variant is not related to the outcome other than via its association with the exposure ^13^. Genetic variants are less likely to be confounded by the socioeconomic and environmental factors that might bias causal estimates in conventional multivariable regression ^14^, but may be biased by violation of their assumptions due to weak or irrelevant instruments, population stratification (causing confounding of the genetic instrument-outcome association) and a path from the genetic instrument to CHD not mediated by the exposure, for example via horizontal pleiotropy or fetal genotype ^15^. Triangulating results from negative control and MR analyses, whereby the key sources of bias differ can help improve the causal understanding of maternal risk factors on CHDs ^16^. Consistent results from both would increase confidence that the relationship is causal. The recent acquisition of genotype information on a large number of maternal-offspring dyads means that we now have relevant data to further test the potential effects of BMI, smoking and alcohol with a complementary method to those used previous. The objective of this study was therefore to explore associations between genetically predicted maternal BMI, smoking and alcohol on offspring CHD using Mendelian randomisation.

## Methods

This study is reported using the Strengthening the Reporting of Observational studies in Epidemiology using Mendelian randomisation (STROBE-MR) guidelines (see Supplementary File: STROBE-MR Checklist)^17,18^.

### Inclusion criteria and participating cohorts

To be eligible for inclusion in this study, cohorts and participants were required to have genome-wide data in mothers and CHD data in the offspring. From previous work with large consortia, including MR-PREG ^19^ and LifeCycle ^20^, we identified three cohorts meeting these criteria: The Avon Longitudinal Study of Parents and Children (ALSPAC), Born in Bradford cohort (BiB), and the Norwegian Mother, Father and Child Cohort Study (MoBa). ALSPAC is a UK prospective birth cohort study which was devised to investigate the environmental and genetic factors of health and development ^21–23^. ALSPAC enrolled pregnant women who resided in and around the city of Bristol in the Southwest of England and had an expected delivery date between April 1, 1991 and December 31, 1992. The enrolled cohort included 15,247 pregnancies resulting in 14,775 live born babies. Ethical approval was obtained from the ALSPAC Ethics and Law Committee and the Local Research Ethics Committees. The study website contains details of all the data that are available through a fully searchable data dictionary and variable search tool. BiB is a population-based prospective birth cohort including 12,453 women across 13,776 pregnancies who were recruited at their oral glucose tolerance test at approximately 26–28 weeks’ gestation ^24^. Eligible women had an expected delivery between March 2007 and December 2010. MoBa is a nationwide, pregnancy cohort comprising family triads (mother-father-offspring) who are followed longitudinally. All pregnant women in Norway who were able to read Norwegian were eligible for participation. The first child was born in October 1999 and the last in July 2009 ^25,26^. One singleton pregnancy per mother in each cohort were included in analyses. **Figure 1** shows the inclusion of participants, after excluding those with missing maternal genotype data and those that did not pass genetic quality control (QC). A total of 38,662 mother-offspring pairs contributed to the main analyses and 28,485 to the adjusted (for fetal genotype) analyses.

**Figure 1.**
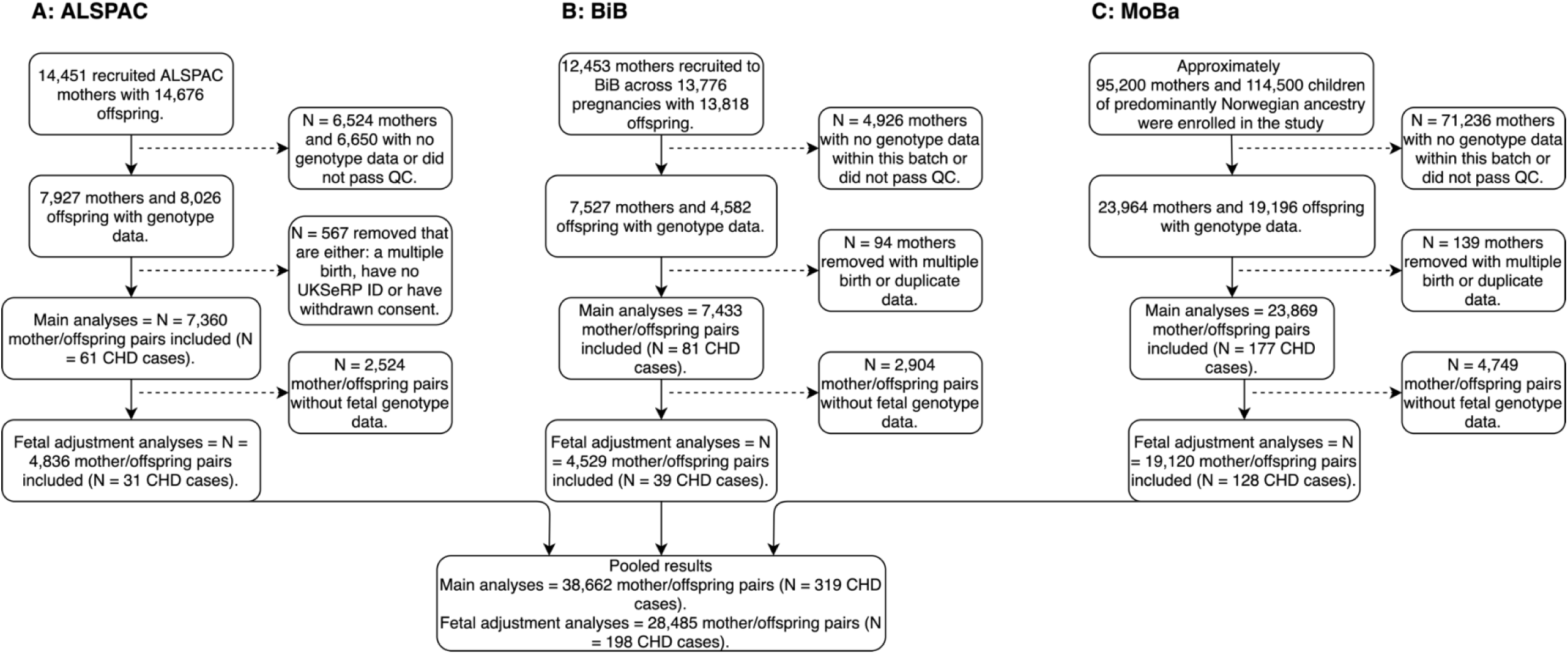
An overview of included cohorts and selection of study participants. Abbreviations: ALSPAC, Avon Longitudinal Study of Parents and Children; BiB, Born in Bradford; MoBa, Norwegian Mother, Father and Child Cohort; QC, quality control; UKSeRP, the secure research platform containing CHD data for ALSPAC; CHD, congenital heart disease; GWAS, genome-wise association study

### Genetic data

#### Genotyping in each cohort

ALSPAC mothers were genotyped using Illumina human660K quad single nucleotide polymorphism (SNP) chip, and ALSPAC children were genotyped using Illumina HumanHap550 quad genome-wide SNP genotyping platform. Genotype data for both ALSPAC mothers and children were imputed against the Haplotype Reference Consortium v1.1 reference panel, after performing the QC procedure (minor allele frequency (MAF) ≥1%, a call rate ≥95%, in Hardy-Weinberg equilibrium (HWE), correct sex assignment, no evidence of cryptic relatedness, and of European descent). The samples of the BiB cohort (mothers and offspring) were processed on three different type of Illumina chips: HumanCoreExome12v1.0, HumanCoreExome12v1.1 and HumanCoreExome24v1.0. Genotype data were imputed against UK10K + 1000 Genomes reference panel, after a similar QC procedure (a call rate ≥99.5%, correct sex assignment, no evidence of cryptic relatedness, correct ethnicity assignment). In MoBa, blood samples were obtained from both parents during pregnancy and from mothers and children (umbilical cord) at birth ^27^. Genotyping has had to rely on several projects - each contributing with resources to genotype subsets of MoBa over the last decade. The data used in the present study was derived from a cohort of genotypes samples from four MoBa batches. The MoBa genetics QC procedure involved MAF ≥1%, a call rate ≥95%, in HWE, correct sex assignment, and no evidence of cryptic relatedness. Further details of the genotyping methods for each cohort are provided in the Supplementary Material (Text S1).

#### GWAS data and SNP selection

We selected SNPs from the largest and most relevant GWAS of European ancestry participants for each exposure (further information for each GWAS shown in Table S1). Selected SNPs were those with a p-value below a p-value threshold used to indicate genome-wide significance after accounting for multiple testing. Of those reaching this threshold we ensured that we only took forward independent SNPs to create the GRSs. This was done either by methods used in the GWAS or by applying our own criteria if the GWAS did not report independent SNP associations. For BMI, there were 941 near-independent SNPs in a combined GWAS of ∼700,000 individuals as reported in Yengo et al ^28^ (near-independent SNPs defined as SNPs with a P < 1×10^−8^ after a conditional and joint multiple SNP analysis to take into account linkage disequilibrium (LD) between SNPs at a given locus). For smoking analyses, there were 126 independent SNPs (genome-wide significant (p<5×10^−8^) SNPs that achieved independence at LD r^2^ = 0.001 and a distance of 10,000 kb). The study was a GWAS of a lifetime smoking index (which combined smoking initiation, duration, heaviness and cessation), conducted in a sample of 462,690 current, former and never smokers in UK Biobank ^29^. A GRS based on the lifetime smoking GWAS has previously been shown to be associated with smoking behaviours during pregnancy in the ALSPAC cohort ^30^. For the alcohol weighted GRS, there were 99 conditionally independent SNPs (P<5×10^−8^), measured as number of alcoholic drinks per week ^31^. This GRS has also previously been shown to be associated with alcohol consumption during pregnancy as well as the general population ^32^. The ALSPAC cohort was included within the original GWAS for alcohol by Liu et al, accounting for 8,913 participants out of a total sample size of 941,280 (0.9%). Previous work has suggested any bias introduced by this level of overlap would be minimal ^33^. Furthermore, a recent study explored this by excluding ALSPAC from the summary statistics and results were unbiased and largely unchanged ^32^. Therefore, we proceeded using the full summary data for generating the alcohol GRS. All GRSs were generated using summary GWAS data that was derived in both men and women. We were unable to obtain female-specific summary data for these GWAS data. However, we performed checks to ensure the GRSs are robustly associated with the maternal exposure during pregnancy.

#### Genetic risk score generation

Weighted GRSs were calculated for BMI, smoking and alcohol consumption by adding up the number of risk factor increasing alleles among the selected SNPs after weighting each SNP by its effect on the corresponding risk factor:

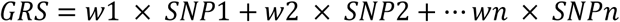

where w is the weight (i.e., the beta-coefficient for the SNP-exposure association reported from the published GWAS) and SNP is the genotype dosage of exposure-increasing alleles at that locus (i.e., 0, 1, or 2 exposure-increasing alleles). Selected SNPs were extracted from the imputed genotype data in dosage format using QCTOOL (v2.0) and VCF tools (v 0.1.12b) in ALSPAC and BiB, respectively. PLINK (v1.9) was then used to construct the GRS for each exposure coded so that an increased score associated with increased exposure. In MoBa, we constructed the GRSs from the QC’d data in PLINK format. Further information on GRS construction for each cohort is shown in Text S2.

### Phenotype data

#### CHD data

In the ALSPAC cohort, cases were obtained from a range of data sources, including health record linkage and questionnaire data up until age 25 following European surveillance of congenital anomalies (EUROCAT) guidelines ^34^. In BiB, cases were identified from either the Yorkshire and Humber congenital anomaly register database, which will tend to pick up most cases that were diagnosed antenatally and in the early postnatal period of life, and through linkage to primary care (up until aged 5), which will have picked up any additional cases, in particular those that might have been less severe and not identified antenatally/in early life ^35^. All these cases were confirmed postnatally and were assigned international classification of disease Version 10 (ICD-10) codes. ICD-10 codes were used to assign CHD cases according to EUROCAT guidelines. In MoBa, information on whether a child had a CHD or not (yes/no) was obtained through linkage to the Medical Birth Registry of Norway (MBRN). All maternity units in Norway must notify births to the MBRN, and information on malformations are reported to the registry up to 12 months postpartum ^36^. Further details on defining CHDs including ICD codes used (in ALSPAC and BiB) are shown in Text S3 and Table S2.

#### Pregnancy phenotype data

As noted above, the SNP selection and weights for the GRS were taken from GWAS in women and men ^28,29,31^. To determine their relevance in women during pregnancy we examined the associations of the GRS with pre/early pregnancy BMI, and pregnancy smoking and alcohol consumption in each cohort. In ALSPAC and MoBa, pre-pregnancy weight and height were self-reported during the first pregnancy questionnaires. In BiB, weight and height were measured at the recruitment assessment. As the timing of questions and the details requested for smoking during pregnancy differed across the three cohorts ^37–39^ we were only able to generate a simple binary variable of any smoking in pregnancy versus none. There was insufficient data and/or power across the cohorts to be able to generate a measure of smoking heaviness in pregnancy. As with smoking, the aim for alcohol was to determine whether the GRS was robustly associated with drinking status during pregnancy. We used questionnaire data in each cohort and used binary variables (yes/no) for whether women consumed any alcohol during pregnancy or not. Further details regarding these phenotype data, including questionnaire information are described in Text S4.

### Statistical analysis

Analyses were performed in R version 4.0.2 (R Foundation for Statistical Computing, Vienna, Austria). A pre-specified analysis plan was uploaded to the Open Science Framework ^40^. We undertook MR in each of the 3 cohorts, including all ALSPAC, BiB and MoBa participants, with maternal genetic data and offspring CHD data. Logistic regression was used to estimate the odds ratio (OR) of CHD per 1 standard deviation (SD) higher GRS, with adjustment for the first 10 genetic principal components (PCs) with additional adjustment for genetic chip, genetic batch, and imputation batch in MoBa.

The key assumptions of MR are: (i) relevance assumption, (ii) independence assumption and (iii) exclusion restriction. To explore the relevance of the GRS to each exposure in pregnancy, we undertook linear (BMI) and logistic (smoking and alcohol) regression to derive the difference in mean BMI and OR of pregnancy smoking and pregnancy alcohol consumption per 1SD higher GRS in each cohort. For BMI, instrument strength was assessed with F-statistics and R^2^. For smoking and alcohol, instrument strength was assessed using the area under the ROC curve and pseudo-R^2^ by the Nagelkerke method ^41^. To minimise the potential for confounding of the GRS-CHD association due to population stratification, we adjusted for the first 10 ancestry-informative PCs ^42^. We also repeated the MR analyses without the inclusion of BiB, given that BiB has a unique ethnic structure of South Asians and White Europeans. To explore horizontal pleiotropy, we checked the association of GRSs with known risk factors for CHD that we had data on (education, parity and diabetes) using linear or logistic regression. Information on how these risk factors were assessed in each cohort is provided in the Supplementary Material (Text S4). If any of the GRSs were associated with another risk factor, we considered that a potential pleiotropic effect. We then performed multivariable MR (MVMR) analyses if GWAS data for the potential pleiotropic variable was available ^43^. Methods for these GRSs and the rationale for selecting these risk factors are described in Text S5. In sensitivity analyses to explore potential bias via fetal genotype we repeated the PC (and batch) adjusted GRS-CHD association in the subsample of participants with fetal genome wide data (**Figure 1**) and then compared those results with the same associations additionally adjusted for the fetal GRS. GRS-CHD association results were pooled using a random effects meta-analysis for all three cohorts and fixed-effect meta-analyses when excluding BiB in sensitivity analyses (i.e., ALSPAC and MoBa). Between study heterogeneity was assessed using the Cochrane Q-statistic and I^2^ ^44^.

## Results

### Participant characteristics

Analyses included 38,662 mother-offspring pairs, of which 319 offspring had CHD (**Figure 1**). The distributions of offspring and maternal characteristics for these analyses in ALSPAC, BiB and MoBa are displayed in **Table 1**. The prevalence of any CHD, mean maternal age and pre-/early-pregnancy BMI were similar in the three cohorts. Women in ALSPAC were more likely to smoke during pregnancy in comparison to those in BiB and MoBa although, the overall prevalence in BiB masks marked differences between the two largest ancestral groups, with 3.4% of South Asian women reporting smoking during pregnancy compared to 34% of White European women. Women in ALSPAC and BiB were more likely to consume alcohol compared to those in MoBa, although, in BiB, there are limited data available on alcohol consumption with very few South Asians responding to questions relating to alcohol in questionnaires.

**Table 1.** Participant characteristics for the 3 studies included in Mendelian randomization analyses.

| Characteristic | Category | ALSPAC (N = 7,360) | BiB (N = 7,433) | MoBa (N = 23,869) |
| --- | --- | --- | --- | --- |
| **Offspring** |  |  |  |  |
| CHD | Yes | 61 (0.8) | 81 (1.1) | 177 (0.7) |
| Sex | Male | 3,703 (50.3) | 3,818 (51.4) | 12,139 (50.9) |
|  | Female | 3,657 (49.7) | 3,615 (48.6) | 11,704 (49.0) |
| **Maternal** |  |  |  |  |
| Age, years |  | 29.2 (4.6) | 27.4 (5.6) | 30.1 (4.5) |
| Parity | Primiparous | 3,257 (46.6) | 2,963 (40.1) | 11,288 (47.3) |
| BMI, kg/m<sup>2</sup> |  | 22.5 (4.2) | 26.2 (5.7) | 24.1 (4.3) |
| Ethnicity | White European | 7,360 (100.0) <sup>a</sup> | 3,084 (42.6) | 23,869 (100.0) <sup>b</sup> |
|  | South Asian | - | 3,503 (48.4) | - |
|  | Other | - | 656 (9.1) | - |
| Any smoking during pregnancy | Yes | 1,679 (26.1) | 1,175 (18.1) | 1,814 (8.6) |
| Any alcohol during pregnancy | Yes | 4,866 (79.9) | 1,040 (49.3) | 6,209 (31.5) |
| Data are means ± SD or n (%) unless stated. % are based on data available (data were not complete).<br><sup>a</sup> All non-white European women with ethnicity data were not included in the analysis.<br><sup>b</sup> Individuals of non-European ancestries were removed based on principal component analysis<br>Abbreviations: BiB, Born in Bradford; ALSPAC, Avon Longitudinal Study of Parents and Children; MoBa, Norwegian Mother, Father and Child Cohort Study; CHD, congenital heart disease; BMI, body mass index; kg, kilograms; m, meters. |  |  |  |  |

### MR results

There were similar statistically positive associations of the BMI GRS with pre-pregnancy BMI and the smoking GRS with pregnancy smoking in all three cohorts (**Table 2**). The alcohol GRS also associated positively with alcohol consumption during pregnancy in all three cohorts with a somewhat weaker association in BiB and MoBa in comparison to ALSPAC. R^2^ and F-statistics for BMI suggested strong instruments, whereas for smoking and alcohol in particular the AUC suggested possible weak instruments.

**Table 2.** Relevance and strength of the genetic risk scores with exposures in pregnancy.

| Study | N participants | N SNPs in GRS | Coefficient (95% CI)<sup>a</sup> | P-Value | R<sup>2</sup>/ pseudo R<sup>2b</sup> | F statistic <sup>c</sup> | AUC |
| --- | --- | --- | --- | --- | --- | --- | --- |
| **Association of GRS for BMI with pre-/early-pregnancy BMI** |  |  |  |  |  |  |  |
| ALSPAC | 6,253 | 941 | 0.24 (0.21, 0.26) | 1 x 10<sup>-80</sup> | 5.6% | 372 | - |
| BiB | 6,196 | 939 | 0.20 (0.18, 0.23) | 5 x 10<sup>-59</sup> | 4.1% | 268 | - |
| MoBa | 22,533 | 868 | 0.25 (0.24, 0.27) | < 1 x 10<sup>-100</sup> | 6.5% | 1,555 | - |
| **Association of GRS for a lifetime smoking index with any smoking during pregnancy** |  |  |  |  |  |  |  |
| ALSPAC | 6,428 | 126 | 1.27 (1.20, 1.35) | 1 x 10<sup>-16</sup> | 1.6% | - | 0.56 |
| BiB | 6,482 | 126 | 1.36 (1.27, 1.45) | 2 x 10<sup>-20</sup> | 2.2% | - | 0.59 |
| MoBa | 20,981 | 119 | 1.23 (1.17, 1.29) | 7 x 10<sup>-17</sup> | 0.8% | - | 0.56 |
| **Association of GRS for drinks per week with any alcohol consumption in pregnancy** |  |  |  |  |  |  |  |
| ALSPAC | 6,087 | 98 | 1.14 (1.07, 1.21) | 3 x 10<sup>-5</sup> | 0.4% | - | 0.53 |
| BiB | 2,110 | 99 | 1.08 (0.99, 1.18) | 0.09 | 0.2% | - | 0.52 |
| MoBa | 19,737 | 73 | 1.02 (0.99, 1.05) | 0.13 | 0.02% | - | 0.51 |
| MoBa sensitivity <sup>d</sup> | 19,737 | 73 | 1.06 (1.01, 1.10) | 0.01 | 0.07% | - | 0.52 |

<sup>a</sup> Effect estimates (coefficient) are difference in mean (BMI) or odds ratio (smoking or drinking yes/no during pregnancy) per SD increase in genetic risk score.
<sup>b</sup> for the binary outcomes (smoking and alcohol) pseudo-R<sup>2</sup> are presented
<sup>c</sup> for BMI F-statistic is presented; for binary outcomes (smoking and alcohol) AUC is presented.
<sup>d</sup> in MoBa 7,356/23,784 consumed any alcohol during pregnancy. However, 4,754 of the 7,356 consumed alcohol “less than once per month” based on the questionnaire data. In the sensitivity analysis shown above, we re-coded the variable so that those that consumed alcohol less than once per month were classed as non-drinkers (N.B. due to the small numbers in each individual category, we were not able to analyze these separately). This was performed as an additional check to ensure the GRS was associated with pregnancy alcohol consumption in MoBa.
Abbreviations: SNP, single nucleotide polymorphism; GRS, genetic risk score; CI, confidence interval; AUC, area under the curve; ALSPAC, Avon Longitudinal Study of Parents and Children; BiB, Born in Bradford; MoBa, Norwegian Mother, Father and Child Cohort.

The MR effects in each study and pooled across studies of each exposure and offspring CHDs are shown in **Figure 2**. For associations of the maternal GRS for BMI and offspring CHD, the pooled OR was close to the null value of 1, but with wide confidence intervals (CI) consistent with 10% lower to 13% higher odds (OR (95% CI) per 1SD higher GRS: 1.01 (0.90, 1.13), with no statistical evidence of between study heterogeneity (**Figure 2A**). When excluding BiB from these analyses, the pooled point estimate increased slightly (OR: 1.06 (0.93, 1.20); Figure S1B Supplementary Material). The BMI GRS associated with smoking, education, and diabetes across all three cohorts (Table S3). Results were unchanged in MVMR models including GRSs for education and smoking (Figures S1C & S1D). 28,485 participants with 198 CHD cases had data on fetal as well as maternal genotype. When the main maternal GRS association was undertaken in this subgroup, the result attenuated (OR: 0.85 (0.60, 1.20)). With additional adjustment for fetal genotype, the result attenuated further in comparison to the unadjusted result in the same subpopulation (OR: 0.81 (0.62, 1.06)). In subgroup analyses for those with fetal genotype data excluding BiB, the pooled results were more consistent and closer to the null (Figures S1E-S1H).

**Figure 2.**
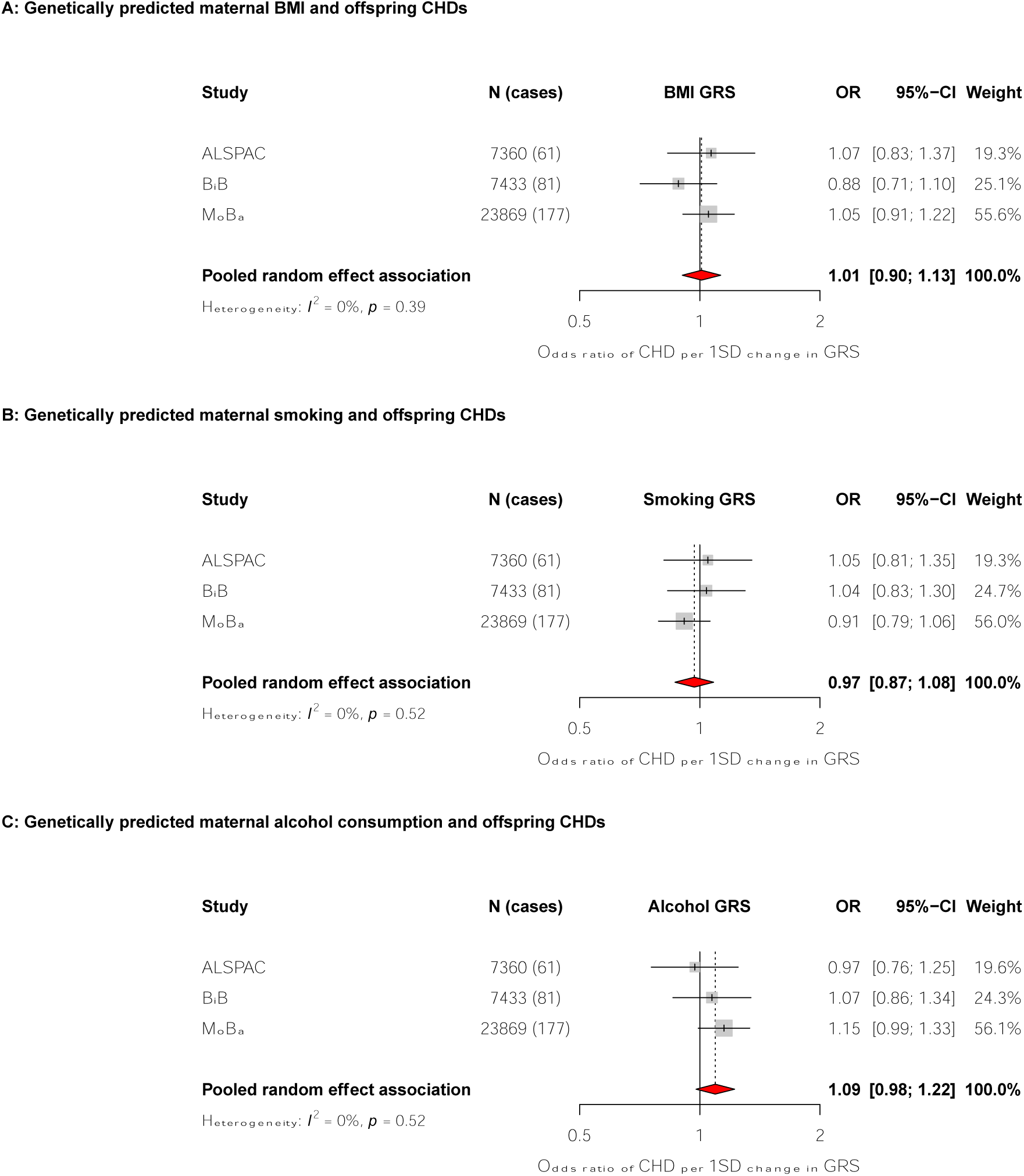
Forest plots showing the mendelian randomisation results for genetically predicted maternal body mass index (Panel A), smoking (GRS of a lifetime smoking index: Panel B), and alcohol consumption (GRS of drinks per week: Panel C) with offspring congenital heart disease. Odds ratios (ORs) of CHD for a 1SD difference in maternal GRS in each study and pooled across studies using random effects meta-analysis. Adjusted for top 10 genetic principal components in all cohorts with additional adjustment for genetic chip, genetic batch, and imputation batch in MoBa. Abbreviations: ALSPAC, Avon Longitudinal Study of Parents and Children; BiB, Born in Bradford; MoBa, Norwegian Mother, Father and Child Cohort Study; BMI, body mass index; CI, confidence interval; CHD, congenital heart disease; SD, standard deviation; GRS, genetic risk score.

The maternal GRS for maternal lifetime smoking index had a pooled OR close to the null, but with wide confidence intervals (OR (95%CI) per 1SD higher GRS: 0.97 (0.87, 1.08), with no statistical evidence of between study heterogeneity (**Figure 2B)**. The smoking GRS associated with BMI and education across the cohorts (Table S4). Results were consistent and materially unchanged in additional analyses when removing BiB (Figure S2B), in MVMR analyses adjusting for education or BMI (Figures S2C & S2D) and in the subgroup analyses with and without adjustment for fetal genotype (Figures S2E-S2H).

There was a positive association between a maternal GRS for alcoholic drinks per week and offspring CHD that was stronger than equivalent results for maternal BMI and smoking (pooled OR: 1.09 (0.98, 1.22)) (**Figure 2C**), with strongest associations found in MoBa. In analyses excluding BiB, the pooled estimated was consistent with main analyses (OR: 1.10 (0.97, 1.25)). The alcohol GRS showed consistent association with smoking across the cohorts (Table S5). The positive association remained in MVMR analyses adjusting for a GRS of smoking (Figure S3C) and in analyses adjusting for offspring genotype (Figures S3D-S3G), although, results were largely being driven by the MoBa cohort.

## Discussion

Using MR across three birth cohorts, we found no strong evidence for an effect of genetically predicted maternal BMI or smoking on risk of offspring CHD but did find evidence of a potential effect of genetically predicted greater alcohol consumption on offspring CHD. However, for all three exposures, confidence intervals were wide and the pseudo R^2^ and AUC suggested potential weak instrument bias for alcohol and smoking. Weak instruments in this study would be expected to bias results toward the confounded association. Weak instruments and imprecise associations also limit clear interpretation of our sensitivity analyses to explore bias due to GRS influencing CHDs via other paths independently of the exposure of interest. We tried to identify all cohorts with maternal genetic data and offspring, CHD measures and to the best of our knowledge this is first MR study of these maternal exposures on offspring CHD risk. Despite the relatively large sample, our inconclusive findings highlight the importance of existing and new cohorts, many of which have genomic data, linking to health care records to obtain information on CHDs and other rare outcomes, for example through electronic health records.

This MR study complements our previous negative paternal control study ^10^. Our MR analyses of BMI are consistent with our previous negative control study, in suggesting that higher maternal BMI may not causally influence offspring CHD and that previous multivariable regression analyses ^5,7^ were likely confounded. We have not clearly replicated our previous result for smoking, which suggested an increased risk of offspring CHD in women who smoked in pregnancy. However, as noted above our imprecise MR results do not rule out an effect, and future larger MR studies are important. Due to the lack of information on alcohol consumption around the time of their partners pregnancy, previous analyses using a negative control design were inconclusive ^10^. Recent meta-analyses found consistent modest increases in risk of offspring CHD in mothers reporting alcohol consumption in pregnancy, however, many of the included studies did not adjust for confounders ^9,45^, meaning that it is difficult to determine whether the association is a result of alcohol or other characteristics that are related to alcohol and offspring CHDs. The possible effect of alcohol consumption on CHD seen here in MR analyses would benefit from exploration using different analysis methods (e.g. well powered negative controls, cross context analyses ^16^ and larger MR studies).

There are several strengths of the current study. We attempted to identify all studies with relevant data knowing that MR is statistically inefficient, and CHD is a rare outcome. We explored potential bias due to other paths from the GRS to CHDs by examining associations of each GRS with the other two exposures and with other risk factors that might influence CHD and undertook multivariable MR where such associations were found. We also adjusted for offspring genotype in a subsample of the pooled data cohort, which is important in attempting to separate the influence of a path from GRS to CHDs via fetal genotype rather than solely from the mothers exposure ^15^.

The key limitation of this study is that despite a relatively large sample size (N = 38,662) the effect estimates were imprecise due to CHD being a rare condition. Furthermore, the GRS for smoking and alcohol may have been biased by the weak instruments. These limitations importantly contribute to our main and sensitivity analysis results. We were not able to clearly differentiate between horizontal (i.e., where smoking is the main exposure of interest, and an association of the smoking GRS with BMI reflects an independent path) and vertical pleiotropy (i.e., where the relation of the smoking GRS with BMI is downstream of the GRS effect on smoking). We want to adjust for horizontal pleiotropy but not vertical pleiotropy, as the latter would be adjusting away part of the mechanism by which, for example smoking might influence CHD. Adjustment for offspring genotype was only possible in a subsample of the main analysis, making results more imprecise and prone to selection bias. However, it is encouraging that our results do not notably differ in these analyses. MR results may be biased by population stratification confounding. We tried to mitigate against that by adjusting for ancestry PCs and exploring consistency of the main results with results removing BiB. Largely these were consistent but even more imprecise. Whilst we included all participants, including those from non-European ancestries, both MoBa (the largest contributing study) and ALSPAC participants are mostly of White European origin, and the GWAS data used to construct the GRSs were restricted to European participants. Therefore, our results may not generalise to other populations. We were only able to explore associations of GRS with any CHD (analyses of subtypes would have been very imprecise) and therefore could have missed potential effects of these exposures on specific CHD subtypes. The MoBa cohort only had cases diagnosed antenatally or around the time of birth (first year of life) which would increase the chances of outcome misclassification by assigning CHD cases which were diagnosed later in life as non-CHD cases. This misclassification is likely to be random with respect to the GRS (i.e., later age at offspring diagnosis could not influence mothers genotype) and would be expected to bias results towards the null, meaning we may have missed some associations.

Identifying modifiable causal risk factors for the development of CHD is important for developing preventive interventions to reduce the risk of CHDs. Improvements in surgery over the last two decades mean that most patients with CHD now live into adulthood. Nonetheless prevention remains important. Many patients require repeat procedures through childhood and adolescence to accommodate their growth, which produces a burden on them, their family and society. Despite trying to identify all relevant studies our results are inconclusive. They highlight the need for more data with maternal genetic and offspring CHD data. We think this is possible over the coming years as running GWAS is relatively cheap, and most cohort studies increasingly have these data. The following could considerably increase the sample size for MR in this field and result in key advances in preventing CHDs: (i) Add data on CHDs through electronic record linkage to existing cohorts; this was done recently in ALSPAC nearly 30 years after the original pregnancies ^34^. Many of the cohorts that we considered for inclusion had genetic data but no information on CHD (or other congenital anomalies). (ii) Linkage to electronic records should be regularly updated at least until early adulthood so that cases that are diagnosed later in life are also captured ^34,35^. (iii) Ensure new cohorts, particularly large birth/pregnancy cohorts or those with the potential to prospectively collect data during pregnancy (such as the planned UK Our Future Health (https://ourfuturehealth.org.uk/about-us/) gain consent to collect health data on CHDs (and other congenital anomalies). (iv) Continue to update the cohorts used in this study and update our results. For example, there are plans to continue running GWAS assays on mothers, fathers and offspring in MoBa who are currently not genotyped which will more than double the sample available in that study. (v) To the best of our knowledge, there are currently no publicly available GWAS summary statistics for CHD. To date, the largest GWAS for CHD in a European population included ∼4,000 cases ^46^. As these GWAS continue to grow, significant data sharing and collaboration will be required, which could then pave way for large-scale two-sample MR studies to explore maternal risk factors for CHDs.

The analysis steps taken in this paper aimed to explore the presence of a causal effect of maternal BMI, smoking and alcohol on offspring CHDs. In summary, we found no robust evidence of an effect for maternal genetically determined BMI or smoking on offspring CHD. We did observe a weak relationship between genetically predicted maternal alcohol intake on offspring CHDs, but this may be explained by weak instrument bias. Despite a large sample size, our results produced imprecise estimates. We have highlighted the need for future larger studies that employ a range of causal methods to further interrogate maternal gestational risk factors for offspring CHDs.

## Supporting information

STROBE-MR Checklist

## Data Availability

The ALSPAC data management plan (http://www.bristol.ac.uk/alspac/researchers/data-access/documents/alspac-data-management-plan.pdf) describes in detail the policy on data sharing, which is through a system of managed open access. Scientists are encouraged to make use of the BiB study data, which are available through a system of managed open access. Please note that the study website contains details of all the data that is available through a fully searchable data dictionary and variable search tool and reference the following webpage:
http://www.bristol.ac.uk/alspac/researchers/our-data/. Before you contact BiB study, please make sure you have read the Guidance for Collaborators: https://borninbradford.nhs.uk/research/guidance-for-collaborators/). MoBa data are used by researchers and research groups at both the Norwegian Institute of Public Health and other research institutions nationally and internationally. The research must adhere to the aims of MoBa and the participants given consent. All use of data and biological material from MoBa is subject to Norwegian legislation. More information can be found on the study website (https://www.fhi.no/en/studies/moba/for-forskere-artikler/research-and-data-access/).

## List of abbreviations

CHD: congenital heart disease
BMI: body mass index
MR: Mendelian randomization
MoBa: Norwegian Mother, Father and Child Cohort Study
GRS: genetic risk score
GWAS: genome-wide association study
ALSPAC: Avon Longitudinal Study of Parents and Children
BiB: Born in Bradford
QC: quality control
SNP: single nucleotide polymorphism
MAF: minor allele frequency
HWE: Hardy-Weinberg equilibrium
LD: linkage disequilibrium
EUROCAT: European surveillance of congenital anomalies
MBRN: medical birth registry of Norway
ICD-10: international classification of disease version 10
STROBE-MR: strengthening the reporting of observational studies in epidemiology using mendelian randomization
OR: odds ratio
SD: standard deviation
PC: principal component
CI: confidence interval
MVMR: multivariable Mendelian randomization

## Declarations

### Ethics approval and consent to participate

Ethical approval for ALSPAC was obtained from the ALSPAC Law and Ethics committee and local research ethics committees (NHS Haydock REC: 10/H1010/70). Informed consent for the use of data collected via questionnaires and clinics was obtained from participants following the recommendations of the ALSPAC Ethics and Law Committee at the time. At age 18, study children were sent ‘fair processing’ materials describing ALSPAC’s intended use of their health and administrative records and were given clear means to consent or object via a written form. Data were not extracted for participants who objected, or who were not sent fair processing materials. For BiB, Ethics approval has been obtained for the main platform study and all of the individual sub studies from the Bradford Research Ethics Committee. Written consent was obtained from all participants. The establishment of MoBa and initial data collection was based on a license from the Norwegian Data Protection Agency and approval from The Regional Committees for Medical and Health Research Ethics. The MoBa cohort is now based on regulations related to the Norwegian Health Registry Act.

### Availability of data and materials

The ALSPAC data management plan (http://www.bristol.ac.uk/alspac/researchers/data-access/documents/alspac-data-management-plan.pdf) describes in detail the policy on data sharing, which is through a system of managed open access. Scientists are encouraged to make use of the BiB study data, which are available through a system of managed open access. Please note that the study website contains details of all the data that is available through a fully searchable data dictionary and variable search tool” and reference the following webpage: http://www.bristol.ac.uk/alspac/researchers/our-data/. Before you contact BiB study, please make sure you have read the Guidance for Collaborators: https://borninbradford.nhs.uk/research/guidance-for-collaborators/). MoBa data are used by researchers and research groups at both the Norwegian Institute of Public Health and other research institutions nationally and internationally. The research must adhere to the aims of MoBa and the participants’ given consent. All use of data and biological material from MoBa is subject to Norwegian legislation. More information can be found on the study website (https://www.fhi.no/en/studies/moba/for-forskere-artikler/research-and-data-access/).

## Authors contributions

K.T. and D.A.L. conceived the study. D.A.L., R.W., J.W., T.Y. and M.M. performed the data curation. K.T. conducted the analysis. K.T. and D.A.L. drafted the original version of the manuscript. All authors provided data interpretation, critical review, and commentary to the revised versions of the manuscript. All authors have seen and approved the final versions of this manuscript.

## Acknowledgements

We are grateful to all the families who took part in the ALSPAC (Avon Longitudinal Study of Parents and Children), the midwives for their help in recruiting them, and the whole ALSPAC team, which includes interviewers, computer and laboratory technicians, clerical workers, research scientists, volunteers, managers, receptionists, and nurses. BiB (Born in Bradford) study is only possible because of the enthusiasm and commitment of the children and parents. We are grateful to all the participants, practitioners, and researchers who have made BiB study happen. The Norwegian Mother, Father and Child Cohort Study is supported by the Norwegian Ministry of Health and Care Services and the Ministry of Education and Research. We are grateful to all the participating families in Norway who take part in this on-going cohort study. We thank the Norwegian Institute of Public Health (NIPH) for generating high-quality genomic data. This research is part of the HARVEST collaboration, supported by the Research Council of Norway (#229624). We also thank the NORMENT Centre for providing genotype data, funded by the Research Council of Norway (#223273), Southeast Norway Health Authorities and Stiftelsen Kristian Gerhard Jebsen. We further thank the Center for Diabetes Research, the University of Bergen for providing genotype data and performing quality control and imputation of the data funded by the ERC AdG project SELECTionPREDISPOSED, Stiftelsen Kristian Gerhard Jebsen, Trond Mohn Foundation, the Research Council of Norway, the Novo Nordisk Foundation, the University of Bergen, and the Western Norway Health Authorities.

## References

1. Wang H, Naghavi M, Allen C, Barber RM, Bhutta ZA, Carter A, et al. Global, regional, and national life expectancy, all-cause mortality, and cause-specific mortality for 249 causes of death, 1980–2015: a systematic analysis for the Global Burden of Disease Study 2015. The Lancet [Internet]. 2016 Oct [cited 2020 Feb 20];388(10053):1459–544. Available from: https://linkinghub.elsevier.com/retrieve/pii/S0140673616310121

2. McCracken Courtney, Spector Logan G., Menk Jeremiah S., Knight Jessica H., Vinocur Jeffrey M., Thomas Amanda S., et al. Mortality Following Pediatric Congenital Heart Surgery: An Analysis of the Causes of Death Derived From the National Death Index. Journal of the American Heart Association [Internet]. 2018 Nov 20 [cited 2020 May 7];7(22):e010624. Available from: https://www.ahajournals.org/doi/10.1161/JAHA.118.010624

3. van der Bom T, Zomer AC, Zwinderman AH, Meijboom FJ, Bouma BJ, Mulder BJM. The changing epidemiology of congenital heart disease. Nature Reviews Cardiology [Internet]. 2011 Jan [cited 2020 May 7];8(1):50–60. Available from: https://www.nature.com/articles/nrcardio.2010.166

4. Jenkins KJ, Correa A, Feinstein JA, Botto L, Britt AE, Daniels SR, et al. Noninherited Risk Factors and Congenital Cardiovascular Defects: Current Knowledge: A Scientific Statement From the American Heart Association Council on Cardiovascular Disease in the Young: Endorsed by the American Academy of Pediatrics. Circulation [Internet]. 2007 Jun 12 [cited 2020 Feb 5];115(23):2995–3014. Available from: https://www.ahajournals.org/doi/10.1161/CIRCULATIONAHA.106.183216

5. Liu X, Ding G, Yang W, Feng X, Li Y, Liu H, et al. Maternal Body Mass Index and Risk of Congenital Heart Defects in Infants: A Dose-Response Meta-Analysis. Biomed Res Int. 2019;1315796.

6. Persson M, Cnattingius S, Villamor E, Söderling J, Pasternak B, Stephansson O, et al. Risk of major congenital malformations in relation to maternal overweight and obesity severity: cohort study of 1.2 million singletons. BMJ [Internet]. 2017 Jun 14 [cited 2020 Feb 5];j2563. Available from: http://www.bmj.com/lookup/doi/10.1136/bmj.j2563

7. Persson M, Razaz N, Edstedt Bonamy A-K, Villamor E, Cnattingius S. Maternal Overweight and Obesity and Risk of Congenital Heart Defects. J Am Coll Cardiol. 2019 08;73(1):44–53.

8. Zhao L, Chen L, Yang T, Wang L, Wang T, Zhang S, et al. Parental smoking and the risk of congenital heart defects in offspring: An updated meta-analysis of observational studies. Eur J Prev Cardiol. 2019 Mar 23;27(12):1284–93.

9. Zhang S, Wang L, Yang T, Chen L, Zhao L, Wang T, et al. Parental alcohol consumption and the risk of congenital heart diseases in offspring: An updated systematic review and meta-analysis. Eur J Prev Cardiolog [Internet]. 2020 Mar [cited 2020 Mar 20];27(4):410–21. Available from: http://journals.sagepub.com/doi/10.1177/2047487319874530

10. Taylor K, Elhakeem A, Thorbjørnsrud Nader JL, Yang TC, Isaevska E, Richiardi L, et al. Effect of Maternal Prepregnancy/Early-Pregnancy Body Mass Index and Pregnancy Smoking and Alcohol on Congenital Heart Diseases: A Parental Negative Control Study. JAHA [Internet]. 2021 Jun [cited 2021 Jun 1];10(11). Available from: https://www.ahajournals.org/doi/10.1161/JAHA.120.020051

11. Davey Smith D. Negative Control Exposures in Epidemiologic Studies: Epidemiology [Internet]. 2012 Mar [cited 2020 Mar 20];23(2):350–1. Available from: http://journals.lww.com/00001648-201203000-00027

12. Davey Smith G, Ebrahim S. ‘Mendelian randomization’: can genetic epidemiology contribute to understanding environmental determinants of disease?*. International Journal of Epidemiology [Internet]. 2003 Feb [cited 2020 Nov 13];32(1):1–22. Available from: https://academic.oup.com/ije/article-lookup/doi/10.1093/ije/dyg070

13. Davies NM, Holmes MV, Davey Smith G. Reading Mendelian randomisation studies: a guide, glossary, and checklist for clinicians. BMJ [Internet]. 2018 Jul 12 [cited 2021 Oct 11];k601. Available from: https://www.bmj.com/lookup/doi/10.1136/bmj.k601

14. Smith GD, Lawlor DA, Harbord R, Timpson N, Day I, Ebrahim S. Clustered Environments and Randomized Genes: A Fundamental Distinction between Conventional and Genetic Epidemiology. Cardon L, editor. PLoS Med [Internet]. 2007 Dec 11 [cited 2020 Dec 10];4(12):e352. Available from: https://dx.plos.org/10.1371/journal.pmed.0040352

15. Lawlor DA, Richmond R, Warrington N, McMahon G, Smith GD, Bowden J, et al. Using Mendelian randomization to determine causal effects of maternal pregnancy (intrauterine) exposures on offspring outcomes: Sources of bias and methods for assessing them. Wellcome Open Res [Internet]. 2017 Feb 14 [cited 2020 Nov 11];2:11. Available from: https://wellcomeopenresearch.org/articles/2-11/v1

16. Lawlor DA, Tilling K, Davey Smith G. Triangulation in aetiological epidemiology. Int J Epidemiol [Internet]. 2017 Jan 20 [cited 2020 Mar 27];dyw314. Available from: https://academic.oup.com/ije/article-lookup/doi/10.1093/ije/dyw314

17. Skrivankova VW, Richmond RC, Woolf BAR, Davies NM, Swanson SA, VanderWeele TJ, et al. Strengthening the reporting of observational studies in epidemiology using mendelian randomisation (STROBE-MR): explanation and elaboration. BMJ [Internet]. 2021 Oct 26 [cited 2022 Jan 3];n2233. Available from: https://www.bmj.com/lookup/doi/10.1136/bmj.n2233

18. Skrivankova VW, Richmond RC, Woolf BAR, Yarmolinsky J, Davies NM, Swanson SA, et al. Strengthening the Reporting of Observational Studies in Epidemiology Using Mendelian Randomization: The STROBE-MR Statement. JAMA [Internet]. 2021 Oct 26 [cited 2022 Jan 3];326(16):1614. Available from: https://jamanetwork.com/journals/jama/fullarticle/2785494

19. Yang Q, Borges MC, Sanderson E, Magnus MC, Kilpi F, Collings PJ, et al. Associations of insomnia on pregnancy and perinatal outcomes: Findings from Mendelian randomization and conventional observational studies in up to 356,069 women. medRxiv [Internet]. 2021 Jan 1;2021.10.07.21264689. Available from: http://medrxiv.org/content/early/2021/10/10/2021.10.07.21264689.abstract

20. Jaddoe VWV, Felix JF, Andersen A-MN, Charles M-A, Chatzi L, Corpeleijn E, et al. The LifeCycle Project-EU Child Cohort Network: a federated analysis infrastructure and harmonized data of more than 250,000 children and parents. Eur J Epidemiol [Internet]. 2020 Jul 23 [cited 2020 Jul 27];709–24. Available from: https://doi.org/10.1007/s10654-020-00662-z

21. Boyd A, Golding J, Macleod J, Lawlor DA, Fraser A, Henderson J, et al. Cohort Profile: The ‘Children of the 90s’—the index offspring of the Avon Longitudinal Study of Parents and Children. International Journal of Epidemiology [Internet]. 2013 Feb [cited 2020 Mar 3];42(1):111–27. Available from: https://academic.oup.com/ije/article-lookup/doi/10.1093/ije/dys064

22. Fraser A, Macdonald-Wallis C, Tilling K, Boyd A, Golding J, Davey Smith G, et al. Cohort Profile: the Avon Longitudinal Study of Parents and Children: ALSPAC mothers cohort. Int J Epidemiol. 2013 Feb;42(1):97–110.

23. Northstone K, Lewcock M, Groom A, Boyd A, Macleod J, Timpson N, et al. The Avon Longitudinal Study of Parents and Children (ALSPAC): an update on the enrolled sample of index children in 2019. Wellcome Open Res [Internet]. 2019 Mar 14 [cited 2020 Mar 10];4:51. Available from: https://wellcomeopenresearch.org/articles/4-51/v1

24. Wright J, Small N, Raynor P, Tuffnell D, Bhopal R, Cameron N, et al. Cohort Profile: The Born in Bradford multi-ethnic family cohort study. International Journal of Epidemiology [Internet]. 2013 Aug 1 [cited 2020 Mar 23];42(4):978–91. Available from: https://academic.oup.com/ije/article-lookup/doi/10.1093/ije/dys112

25. Magnus P, Birke C, Vejrup K, Haugan A, Alsaker E, Daltveit AK, et al. Cohort Profile Update: The Norwegian Mother and Child Cohort Study (MoBa). Int J Epidemiol. 2016;45(2):382–8.

26. Magnus P, Irgens LM, Haug K, Nystad W, Skjaerven R, Stoltenberg C, et al. Cohort profile: the Norwegian Mother and Child Cohort Study (MoBa). Int J Epidemiol. 2006 Oct;35(5):1146–50.

27. Rønningen KS, Paltiel L, Meltzer HM, Nordhagen R, Lie KK, Hovengen R, et al. The biobank of the Norwegian Mother and Child Cohort Study: a resource for the next 100 years. Eur J Epidemiol. 2006;21(8):619–25.

28. Yengo L, Sidorenko J, Kemper KE, Zheng Z, Wood AR, Weedon MN, et al. Meta-analysis of genome-wide association studies for height and body mass index in ∼700000 individuals of European ancestry. Hum Mol Genet. 2018 Oct 15;27(20):3641–9.

29. Wootton RE, Richmond RC, Stuijfzand BG, Lawn RB, Sallis HM, Taylor GMJ, et al. Evidence for causal effects of lifetime smoking on risk for depression and schizophrenia: a Mendelian randomisation study. Psychol Med. 2020 Oct;50(14):2435–43.

30. Schellhas L, Haan E, Easey KE, Wootton RE, Sallis HM, Sharp GC, et al. Maternal and child genetic liability for smoking and caffeine consumption and child mental health: an intergenerational genetic risk score analysis in the ALSPAC cohort. Addiction [Internet]. 2021 Nov [cited 2022 Feb 17];116(11):3153–66. Available from: https://onlinelibrary.wiley.com/doi/10.1111/add.15521

31. Liu M, Jiang Y, Wedow R, Li Y, Brazel DM, Chen F, et al. Association studies of up to 1.2 million individuals yield new insights into the genetic etiology of tobacco and alcohol use. Nat Genet [Internet]. 2019 Feb [cited 2021 Jun 3];51(2):237–44. Available from: http://www.nature.com/articles/s41588-018-0307-5

32. Easey KE, Wootton RE, Sallis HM, Haan E, Schellhas L, Munafò MR, et al. Characterization of alcohol polygenic risk scores in the context of mental health outcomes: Within-individual and intergenerational analyses in the Avon Longitudinal Study of Parents and Children. Drug and Alcohol Dependence [Internet]. 2021 Apr [cited 2021 Jun 3];221:108654. Available from: https://linkinghub.elsevier.com/retrieve/pii/S0376871621001496

33. Burgess S, Davies NM, Thompson SG. Bias due to participant overlap in two-sample Mendelian randomization. Genet Epidemiol [Internet]. 2016 Nov [cited 2021 Jun 18];40(7):597–608. Available from: https://onlinelibrary.wiley.com/doi/10.1002/gepi.21998

34. Taylor K, Thomas R, Mumme M, Golding J, Boyd A, Northstone K, et al. Ascertaining and classifying cases of congenital anomalies in the ALSPAC birth cohort. Wellcome Open Res [Internet]. 2020 Oct 6 [cited 2020 Oct 13];5:231. Available from: https://wellcomeopenresearch.org/articles/5-231/v1

35. Bishop C, Small N, Mason D, Corry P, Wright J, Parslow RC, et al. Improving case ascertainment of congenital anomalies: findings from a prospective birth cohort with detailed primary care record linkage. bmjpo [Internet]. 2017 Nov [cited 2020 Mar 4];1(1):e000171. Available from: http://bmjpaedsopen.bmj.com/lookup/doi/10.1136/bmjpo-2017-000171

36. Leirgul E, Fomina T, Brodwall K, Greve G, Holmstrøm H, Vollset SE, et al. Birth prevalence of congenital heart defects in Norway 1994-2009—A nationwide study. American Heart Journal [Internet]. 2014 Dec [cited 2022 Jan 18];168(6):956–64. Available from: https://linkinghub.elsevier.com/retrieve/pii/S0002870314004980

37. Brand JS, Gaillard R, West J, McEachan RRC, Wright J, Voerman E, et al. Associations of maternal quitting, reducing, and continuing smoking during pregnancy with longitudinal fetal growth: Findings from Mendelian randomization and parental negative control studies. PLoS Med [Internet]. 2019 Nov 13 [cited 2020 Jul 1];16(11):e1002972. Available from: https://www.ncbi.nlm.nih.gov/pmc/articles/PMC6853297/

38. Macdonald-Wallis C, Tilling K, Fraser A, Nelson SM, Lawlor DA. Established preeclampsia risk factors are related to patterns of blood pressure change in normal term pregnancy: findings from the Avon Longitudinal Study of Parents and Children. J Hypertens. 2011 Sep;29(9):1703–11.

39. Hernáez Á, Wootton RE, Page CM, Skåra KH, Fraser A, Rogne T, et al. Smoking and subfertility: multivariable regression and Mendelian randomization analyses in the Norwegian Mother, Father and Child Cohort Study. medRxiv [Internet]. 2021 Jan 1;2021.10.25.21265469. Available from: http://medrxiv.org/content/early/2021/10/25/2021.10.25.21265469.abstract

40. Taylor, Kurt. Effect of Maternal Body Mass Index, Smoking and Alcohol on Congenital Heart Diseases: A Mendelian Randomization Study. 2022 [cited 2022 Mar 18]; Available from: https://osf.io/z6g2f/

41. Nagelkerke NJD. A note on a general definition of the coefficient of determination. Biometrika [Internet]. 1991 [cited 2022 Jan 21];78(3):691–2. Available from: https://academic.oup.com/biomet/article-lookup/doi/10.1093/biomet/78.3.691

42. Wang C, Zhan X, Liang L, Abecasis GR, Lin X. Improved ancestry estimation for both genotyping and sequencing data using projection procrustes analysis and genotype imputation. Am J Hum Genet. 2015 Jun 4;96(6):926–37.

43. Burgess S, Thompson SG. Multivariable Mendelian randomization: the use of pleiotropic genetic variants to estimate causal effects. Am J Epidemiol. 2015 Feb 15;181(4):251–60.

44. Higgins JPT, Thompson SG, Deeks JJ, Altman DG. Measuring inconsistency in meta-analyses. BMJ [Internet]. 2003 Sep 6 [cited 2020 May 22];327(7414):557–60. Available from: https://www.ncbi.nlm.nih.gov/pmc/articles/PMC192859/

45. Wen Z, Yu D, Zhang W, Fan C, Hu L, Feng Y, et al. Association between alcohol consumption during pregnancy and risks of congenital heart defects in offspring: meta-analysis of epidemiological observational studies. Ital J Pediatr. 2016 Feb 3;42:12.

46. Lahm H, Jia M, Dreßen M, Wirth FFM, Puluca N, Gilsbach R, et al. Congenital heart disease risk loci identified by genome-wide association study in European patients. Journal of Clinical Investigation [Internet]. 2020 Nov 17 [cited 2020 Nov 23]; Available from: http://www.jci.org/articles/view/141837

