## Supplementary Material for "The effect of maternal BMI, smoking and alcohol on congenital heart diseases: a Mendelian randomization study"

**Text S1.** Genetic data methods.

***The Avon Longitudinal Study of Parents and Children (ALSPAC)***

Mothers were genotyped on Illumina HumanHap660W quad-chip platform by Centre National de Génotypage (Évry, FR). Offspring were genotyped on Illumina HumanHap550 quad-chip platforms by the Wellcome Trust Sanger Institute (Cambridge, UK) and by the Laboratory Corporation of America (Burlington, USA) using support from 23andMe. Standard quality control was applied to SNPs and individuals. Individuals were excluded based on genotype rate (<5%), sex mismatch, high heterozygosity and cryptic relatedness [defined as identity-by-descent (IBD) >0.125]. In order to remove individuals of non-European descent, principal components (PCs) were derived from linkage disequilibrium-pruned SNPs with MAF >0.01 using plink. Individuals laying 5 standard deviations beyond the 1000 Genomes European population PCs 1 and 2 centroid were excluded. SNPs with a minor allele frequency (MAF) <1%, genotyping rate <5% or with a deviation from Hardy–Weinberg disequilibrium (pP << $1\times10{-}6$) were removed from the analysis. Using this QC'd dataset, a list of unrelated mothers was created using an IBD cut-off of 0.05. For imputation, genotypes of ALSPAC mothers and children were combined. Haplotypes were estimated using ShapeIT (v2. r644), which utilises relatedness during phasing. A phased version of the 1000 genomes reference panel (Phase 1, Version 3) was obtained from the Impute2 reference data repository. Imputation was performed using Impute V2.2.2 against the reference panel (all polymorphic SNPs excluding singletons), using all 2186 reference haplotypes (including non-Europeans).

***Born in Bradford (BiB)***

The samples of the BiB cohort (mothers and offspring) were processed on three different type of Illumina chips: HumanCoreExome12v1.0, HumanCoreExome12v1.1 and HumanCoreExome24v1.0. The pre-processing of samples was done separately for the three chips. Problematic samples which had a Call Rate < 0.95 were removed. Poorly performing SNPs determined by a set of quality matrices were zeroed.

*BiB Illumina HumanCoreExome: PLINK and filtering*

GenomeStudio output files were converted to PLINK format and subsequently filtered. SNPs where >=20% of individuals were missing genotype were removed. Individuals with >=10% missing genotypes were removed. A further pass over genotype rate was performed, removing SNPs where over 20% are missing genotype. Following inspection of plink *--missing output*, individuals with > 1% missing genotypes were removed. The final pass over genotype rate, removed SNPs where over 0.5% were missing genotype. The final pass over missingness per individual, removed individuals with over 0.5% missing genotypes.

*Quality control and imputation*

From each of the 3 genotyping sets any individual or SNP missing >3% of their data was dropped and the datasets combined. Genetic duplicates were removed. Reported first degree relatives (mother-child, father-child, child-child siblings) were checked to see if they looked genetically like first degree relatives. If there was no such evidence of this, they were removed. Mother-child discrepancies between phenotype and genotype were removed. People who looked genetically to be clearly South Asian or White British from the principal component analysis (PCA) but had a different ethnicity phenotype were removed. Based on a combination of PCA and reported ethnicity there were two subsets of individuals – white European and south Asian. As a sensitivity analysis, all of the genetic analyses in BiB are repeated after stratifying by these two ethnic groups. SNPs with a minor allele frequency (MAF) <1%, genotyping rate <5% or with a deviation from Hardy–Weinberg disequilibrium (pP << 1×10−6) were removed from the analysis. Imputation was performed for Europeans and South Asians separately, both using the HRC r1.1 as the reference panel. The genotype data was uploaded to the Michigan Imputation Server to perform genotype imputation using Minimac4. Phasing was performed using Eagle v2.4. After imputation, the VCF files were downloaded from the server and BCFtools was used to remove SNPs that were not accurately imputed. Mimimac4 generates a metric (imputation accuracy $R^{-squared}$) for each variant, and variants with estimated imputation accuracy $R^2$ <0.3 were removed.

***The Norwegian Mother, Father and Child Cohort (MoBa)***

Compared to other large biobanks like the UK Biobank, where considerable funding was secured upfront allowing for genotyping their entire cohort in a single effort, genotyping in MoBa have had to rely on several projects - each contributing with resources to genotype subsets of MoBa over the last decade. Consequently, genotyping was performed years apart at different labs using different arrays. We used data from MoBaGenetics 1.0. There is an openly available comprehensive GitHub page that documents all quality control for all releases of genetic data in the MoBa cohort (https://github.com/folkehelseinstituttet/mobagen). In this study we used data from the following batches: NORMENT, ROTTERDAM, TED and HARVEST (initial N = ~98,000). 33,047 individuals were genotyped in the NORMENT sample at deCODE genetics, Reykjavik Iceland (Illumina HumanOmniExpress-24v1.0, Illumina InfiniumOmniExpress-24v1.2, & Illumina Global Screening Array MD v.1.0 + 50k custom OmniExpress overlap content array), 26,680 were genotyped in the ROTTERDAM sample at ERASMUS MC,

Rotterdam, Netherlands (Illumina Global Screening Array MD v.1.0 array), 5215 were sampled in the TED samples at deCODE genetics (Illumina InfiniumOmniExpress-24v1.2), and 32,886 were sampled in the HARVEST sample at Genomics Core Facility, Trondheim, Norway (llumina HumanCoreExome12v1.1 & Illumina HumanCoreExome24v1.0). Below, we describe the methods and QC for the merged dataset used in the present study.

Quality control (QC) and imputation was performed to align with current best-practice QC protocols in human genetics and the family-based pipeline Picopili. The primary software used for the QC was PLINK 1.9 and KING 2.2.5. To identify core subpopulations filtering of was performed for minor allele frequency of 1%, SNP and individual call rate of 95%, and Hardy-Weinberg Equilibrium (HWE) p-value of 0.001. Principle component (PC) analysis with 1000 Genomes phase 1 data was used to identify the European, Asian, and African core subpopulations.

Pre-imputation QC was performed for each of the core subpopulations on the SNP and individual level. QC on a SNP level involved filtering for 0.5% MAF, 95% call rate, HWE p-value 0.000001, discordant in duplicate pairs, association with genotype plate and genotype batch at p-value 0.001. Individual level QC was performed by filtering for heterozygosity outliers Fhet ± 0.2, erroneous sex assignment, known relatedness, cryptic relatedness, identity-by-decent (PI_HAT threshold of 0.15), and PC outliers both with and without 1000 Genomes. Mendel errors were assessed for families with a minimum of one PO duo. Families with more than 5% Mendel errors and SNPs with more than 1% of Mendel errors were removed, while other minor Mendel errors were zeroed out. Batches that were genotyped using the same array were merged (keeping only SNPs present in all batches) and the pre-imputation QC was performed on the merged batches.

Phasing and imputation was performed using the publicly available Haplotype Reference Consortium data. Phasing was performed using SHAPEIT2 with the duoHMM algorithm to incorporate the pedigree information into the haplotype estimates. IMPUTE 4 was then used to perform imputation. Dosage data was then converted to best-guess, hard call genotype data with an imputation quality score (INFO) of 0.8 and default PLINK certainty of 0.9. Post-imputation QC was then performed following the steps outlined in the pre-imputation QC. To ensure the across batch relatedness (both known, such as PO and FS relationships, and unknown, such as sibships within the parent generation) was accounted for in all analyses the three imputation batches were merged, and post-imputation QC was performed on the overall merged dataset. We removed related individuals (cryptic relatedness: IBD >0.05).

**Table S1.** Further information on the genome-wide association studies used to generate genetic risk scores.

| *Primary outcome* | *Data* | *Publication year* | *N* | *Ancestry* | *Imputation reference panel* | *Control for population structure* | *Model* | *Covariables* | *Units* | *PMID* | *Data access* |
|---|---|---|---|---|---|---|---|---|---|---|---|
| Body mass index | UKBB and GIANT | 2018 | ~700,000 | European | HRC imputation reference panel | 10 PCs | Linear mixed model - BOLT-LMM v2.3 | Age, sex, recruitment centre, genotyping batches and 10 PCs | Kg/m$^2$ | 30124842 | Open access. |
| Lifetime smoking – heaviness, duration, initiation | UKBB | 2019 | 462,690 | European | UK 10K reference panel | 10 PCs | Linear mixed model - BOLT-LMM | Genotype chip, sex, 10PCs | Lifetime smoking score (mean = 0.36). | 31689377 | Open access. |
| Alcoholic drinks per week | Large consortium – see paper for all studies. | 2019 | 941,280 | Mostly European or US. | Most studies used HRC imputation reference panel. | 10 PCs. | Linear mixed model with a genetic kinship matrix. | PCs, age, sex, age x sex interaction. | Alcoholic drinks per week | 30643251 | Open access. |

PMID, PubMed ID number; UKBB, UKBioBank; HRC, haplotype reference consortium; PC, principal component.

**Text S2.** Genetic risk score generation.

***ALSPAC***

Selected SNPs were extracted from the imputed genotype data in dosage format using QCTOOL (v2.0). PLINK (v1.9) was then used to construct the GRS for each exposure coded so that an increased score associated with increased exposure.

***BiB***

Selected SNPs were extracted from the imputed genotype data in dosage format using VCF tools (v 0.1.12b). PLINK (v1.9) was then used to construct the GRS for each exposure coded so that an increased score associated with increased exposure.

***MoBa***

In MoBa, we constructed the GRSs from the QC'd data in PLINK format. In MoBa, there were a large proportion of missing SNPs. We subset SNPs included in full GWAS results to SNPs also available in the QC'd MoBa data. From SNPs available in both, independent genome-wide significant associations were identified by clumping in MRBase, specifying r=0.01 and $p<5.0x10^{-8}$ [1]. Subsetting to SNPs available in MoBa and then clumping within these avoids the need for an additional step identifying proxy SNPs. These steps produced a similar number of SNPs for the BMI and smoking GRS in comparison to the GRSs generated in ALSPAC and BiB (941 and 939 in ALSPAC and BiB, respectively vs 868 in MoBa for BMI and 126 in ALSPAC and BiB vs 119 in MoBa for smoking). However, for alcohol, there was significantly less SNPs (98 and 99 in ALSPAC and BiB, respectively vs 37 in MoBa) most likely due to the approach the original GWAS used which was not possible to replicate. Therefore, as an alternative, we used the same summary data as ALSPAC and BiB and used a proxy SNP where available based on $r^2 > 0.8$ using the European reference panel in the LDLink R package [2], which left a total of 73 SNPs.

**Text S3.** Defining congenital heart disease.

***ALSPAC***

Case ascertainment of CAs in the ALSPAC cohort has been described in detail in a recently published data note [29]. Data were combined from multiple sources: NHS records (primary care, paediatric cardiology database, data on fetal deaths and local child health services), midwifery and birth records and maternal self-report via child-based questionnaires. Each source was coded using ICD-10 codes. By combining sources, there would be a greater possibility of capturing all of possible cases within the cohort. The majority of cases of CAs were identified by primary care records (79% for any CA and 68% for any CHD). We included diagnoses made at any age (from birth up until age 25/26). There were no restrictions in cases of CAs in ALSPAC, we included all cases whether live-born or not. However, it is possible that some cases that were terminated earlier in pregnancy were missed due to them never having an NHS number and thus not being identified through record linkage.

***BiB***

In the BiB cohort, there were two separate sources to identify CAs. Both sources were used in this study: (i) CAs up to 5 years of age, identified in GP records by Bishop et al [30] following EUROCAT guidelines. ICD-10 codes were mapped to clinical term (CT)-V3 codes prior to extraction from GP records. (ii) Data extracted from the Yorkshire and Humber CAs register database. Data were ICD-10 coded. All of these were confirmed postnatally. BiB includes data on the birth outcome of each child (live birth, miscarriage, still birth). Therefore, diagnoses were not necessarily restricted to live born children. However, there is the possibility that some women would have terminated the pregnancy after the 12- or 20-week scans which would lead to an under-representation of congenital anomaly cases.

***MoBa***

Information on whether a child had a CHD or not was obtained though linkage to the Medical Birth Registry of Norway (MBRN). All maternity units in Norway must notify births to the MBRN. Further information can be found in the publication by Leirgul et al (https://doi.org/10.1016/j.ahj.2014.07.030). The notification form includes the name and personal identity number of the child and parents, as well as information about maternal health before and during pregnancy, and any complications during pregnancy or at birth, including the presence of any heart defects. The MBRN contains information on all births and pregnancies ended after the 12th week of gestation, including stillbirths and abortions after the 12th week, including on heart defects. Heart defects are registered in the MBRN through notifications from clinical staff identifying these defects at delivery or any hospital in patient treatments occurring immediately after birth until the child is discharged. The medical notification is made at discharge, which can be several months after birth. Details of the notified heart defects, such as specific diagnosis or treatment are not provided. Whilst most of the heart defects would have been diagnosed at birth it is possible that some children were admitted to hospital after delivery for non-specific reasons of for diagnoses that at the time were not considered to be related to a heart defect. Therefore, we considered MoBa diagnoses to have been made between birth and 6 months (few would remain in hospital after this length).

**Table S2.** Subcategories of CHD.

| Category | CHDs included/excl | ICD-10 codes |
|---|---|---|
| All CHDs | Any CHD as defined by EUROCAT*<br>Patent ductus arteriosus with gestational age < 37 weeks not considered a CHD case.<br>Peripheral pulmonary artery stenosis with gestational age < 37weeks not considered as a CHD case. | Q20-Q25, Q260, Q262-Q269** |

* Definitions taken from here: https://eu-rd-platform.jrc.ec.europa.eu/sites/default/files/EUROCAT-Guide-1.4-Section-3.3.pdf
**Q250 and Q256 not a case if isolated and GA<37weeks
Abbreviations: CHD, congenital heart disease; ICD, international classification of disease; EUROCAT, European surveillance of congenital anomalies.

**Text S4.** Describing the pregnancy phenotype data: maternal BMI, smoking, alcohol, education, parity, diabetes separated by each cohort.

**ALSPAC**

For ALSPAC, in the 2nd pregnancy questionnaire (12 weeks' gestation) women were asked to report their pre-pregnancy weight and height and these were used to calculate BMI. No definition of pre-pregnancy was provided in the question. Extracted first antenatal clinic measurements of weight correlated strongly with the women's self-report (Pearson correlation = 0.93).

For smoking, women were asked the number of cigarettes per day during pregnancy in questionnaires at around 18 weeks' and 32 weeks' gestation. Binary variable used any smoking during pregnancy.

For alcohol, women were asked whether they had consumed alcohol during months 1-3 of the pregnancy in a questionnaire administered at around 18 weeks' and 32 weeks' gestation. Women were also asked about how many units they consumed in a questionnaire at 32 weeks' gestation. Binary variable used any alcohol consumption during pregnancy.

Women were asked about their highest educational qualification in a questionnaire administered around 32 weeks' gestation. Education was defined according to the international classification (High: Short cycle tertiary, Bachelor, Masters, Doctoral or equivalent (ISCED-2011: 5-8, ISCED-97: 5-6) Medium: Upper secondary, Post-secondary non- tertiary (ISCED-2011: 3-4, ISCED-97: 3-4) Low: No education; early childhood; pre-primary; primary; lower secondary or second stage of basic education). A binary variable was used (yes = medium or high education, no = low education).

For parity previous stillbirths were included and abortions excluded. Women were asked about previous children in a questionnaire administered around 32 weeks' gestation. A binary variable was used signifying multiparous and nulliparous women.

For diabetes, women were asked about existing diabetes and pregnancy diabetes using pregnancy questionnaires. Binary variable used any diabetes yes/no.

**BiB**

For BiB, weight and height (unshod and in light clothing and following a standard protocol) were measured at the recruitment assessment. As women were recruited at the oral glucose tolerance test (26-

28 weeks of gestation for the majority) this would not provide an accurate measure of pre-/early-pregnancy weight, as it would include fetal and amniotic weight and pregnancy related weight gain. All measurements of weight from all antenatal clinics were extracted from the obstetric records and pre-/early-pregnancy BMI was calculated using weight from the first antenatal clinic (median 12 weeks' gestation) and height at recruitment (26-28 weeks' gestation).

Women were asked number of cigarettes per day during pregnancy in the first questionnaire (26-28 weeks' gestation). Binary variable used any smoking during pregnancy.

Women were asked whether they consumed alcohol during the first 3 months of pregnancy.

Women were asked about their highest educational qualification in the recruitment questionnaire. Education was defined according to the international classification (High: Short cycle tertiary, Bachelor, Masters, Doctoral or equivalent (ISCED-2011: 5-8, ISCED-97: 5-6) Medium: Upper secondary, Post-secondary non- tertiary (ISCED-2011: 3-4, ISCED-97: 3-4) Low: No education; early childhood; pre-primary; primary; lower secondary or second stage of basic education). A binary variable was used (yes = medium or high education, no = low education).

For parity previous stillbirths were included and abortions excluded. Women were asked about previous children in a questionnaire administered at recruitment. A binary variable was used signifying multiparous and nulliparous women.

For diabetes, women were diagnosed with gestational diabetes based on results from the oral glucose tolerance test at recruitment. This was defined according to modified World Health Organization (WHO) definition used in clinical practice at the time: fasting glucose ≥ 6.1 mmol/L or 2 h post-load glucose ≥ 7.8 mmol/L. We then used questionnaire data that asked about existing diabetes administered at recruitment and defined an "any diabetes" variable.

**MoBa**

For MoBa, pre-pregnancy weight and height were self-reported during the first questionnaire at around 15 weeks' gestation.

During questionnaires administered around 15- and 32-weeks' gestation, women were asked if they smoked now after becoming pregnant. A binary variable was used to signify any smoking during pregnancy.

During the questionnaire administered around 32 weeks' gestation, women were asked about their drinking habits at different time points in the pregnancy. The options were: never, less than once a month, roughly 1-3 times a week, roughly once a week, roughly 2-3 times a week, roughly 4-5 times a week and roughly 6-7 times a week. A binary variable was used to define any drinking during pregnancy (no = those that answered “never”, yes = those that answered anything else). In a sensitivity analysis to check the robustness of the GRS, we defined drinking during pregnancy as: no = those that answered “never” or “less than once a month” and yes = those that answered anything else.

Women were asked about their education in the questionnaire administered around 15 weeks’ gestation. The options were: 1) 9-year secondary school, 2) 1-2 year high school, 3) Vocational high school, 4) 3-year high school general studies, junior college, 5) Regional technical college, 4-year university degree (Bachelor’s degree, nurse, teacher, engineer), 6) University, technical college, more than 4 years (Master’s degree, medical doctor, PhD). We created a binary variable for high education: yes = 5 or 6, no = 1,2,3 or 4.

For parity, women were asked about the number of “previous deliveries” in a questionnaire. A binary variable was used signifying multiparous and nulliparous women.

For diabetes, women were asked about existing diabetes and pregnancy diabetes using pregnancy questionnaires. Binary variable used any diabetes yes/no.

**Text S5.** Genetic risk scores for multivariable Mendelian randomisation (MVMR).

The BMI GRS associated with smoking, education and diabetes across all three cohorts (Table S2). The effect of BMI on diabetes is well established, including from previous MR studies [3–5]. MR evidence suggests that higher education is causally related to lower BMI [6] whereas previous MR analyses show a potential causal effect of higher BMI on initiating smoking and other smoking traits [7,8] as well as smoking causing a reduction in BMI [9]. These findings would suggest that diabetes is a mediating path from BMI to CHD rather than a cause of horizontal pleiotropy, whereas education might be a source of horizontal pleiotropy and smoking, potentially with a bidirectional relationship could be both a horizontal pleiotropic and/or mediating path. Thus, we undertook MVMR adjusting the effects of the BMI GRS by a GRS predicting education (details below), and separately a smoking GRS, in additional analyses of the potential effect of BMI on CHDs, with caution in interpreting any change with adjustment for the smoking GRS.

The smoking GRS associated with BMI and education across the cohorts (Table S3). As discussed above the bidirectional relationship between BMI and smoking make it difficult to decide whether BMI is a potential biasing path, here, between the smoking GRS and CHD or mediates an effect of smoking. We undertook MVMR adjusting for a GRS predicting education (details below), and separately the BMI GRS, in additional analyses of the potential effect of smoking on CHDs.

The alcohol GRS showed consistent association with smoking across the cohorts (Table S4), and we used MVMR to adjust for the smoking GRS, to explore evidence that this might bias any effects of alcohol on CHD. There was evidence of the alcohol GRS relating to smoking and parity in BiB but given the weak statistical evidence and presence only in one of the cohorts we did not explore this further.

*Education GRS*

We used a recent large-scale GWAS on educational attainment [10] (~1.1 million participants, N = 481 independent SNPs in ALSPAC and BiB and 410 independent SNPs in MoBa ($r=0.01$ and $p<5.0 \times 10^{-8}$)). We generated the GRS for education using the same methods as described above (Text S2) and then included the GRS in the MR regression models.

### Exploring associations between the GRSs (BMI, lifetime smoking index, drinks per week) and risk factors for CHDs.

**Table S3.** Exploring associations between the BMI GRS and risk factors for CHDs. We also include the association of the BMI GRS with BMI (also shown in table X within the manuscript) for comparison.

| Risk factor | N | Coefficient (95% CI) [a] | P-value | $R^2$ / Pseudo $R^2$ [b] | F statistic [c] | AUC |
|---|---|---|---|---|---|---|
| **ALSPAC** | | | | | | |
| BMI | 6,253 | 0.24 (0.21, 0.26) | $1 \times 10^{-80}$ | 5.6% | 372 | - |
| Education | 6,806 | 0.87 (0.81, 0.93) | $2 \times 10^{-5}$ | 0.45% | - | 0.54 |
| Parity | 6,982 | 1.03 (0.98, 1.08) | 0.25 | 0.03% | - | 0.51 |
| Diabetes | 6,786 | 1.15 (0.83, 1.60) | 0.17 | 0.16% | - | 0.54 |
| Smoking | 6,428 | 1.14 (1.08, 1.21) | $4 \times 10^{-6}$ | 0.49% | - | 0.54 |
| Alcohol | 6,087 | 0.96 (0.91, 1.03) | 0.27 | 0.03% | - | 0.51 |
| **BiB** | | | | | | |
| BMI | 6,196 | 0.20 (0.18, 0.23) | $5 \times 10^{-59}$ | 4.1% | 268 | - |
| Education | 6,483 | 0.92 (0.88, 0.97) | 0.002 | 0.2% | - | 0.52 |
| Parity | 7,259 | 1.04 (0.99, 1.09) | 0.15 | 0.03% | - | 0.51 |
| Diabetes | 7,133 | 1.10 (1.01, 1.18) | 0.04 | 0.1% | - | 0.52 |
| Smoking | 6,482 | 1.09 (1.02, 1.16) | 0.01 | 0.2% | - | 0.52 |
| Alcohol | 2,110 | 1.07 (0.98, 1.16) | 0.15 | 0.1% | - | 0.52 |
| **MoBa** | | | | | | |
| BMI | 22,533 | 0.25 (0.24, 0.27) | $< 1 \times 10^{-100}$ | 6.5% | 1,555 | - |
| Education | 21,921 | 0.90 (0.87, 0.92) | $3 \times 10^{14}$ | 0.4% | - | 0.53 |
| Parity | 23,869 | 1.00 (0.97, 1.02) | 0.80 | 0.0004% | - | 0.50 |
| Diabetes | 23,869 | 1.24 (1.12, 1.39) | $8 \times 10^{-5}$ | 0.5% | - | 0.56 |
| Smoking | 20,981 | 1.18 (1.12, 1.24) | $2 \times 10^{-11}$ | 0.5% | - | 0.55 |
| Alcohol | 19,737 | 0.97 (0.94, 1.00) | 0.03 | 0.03% | - | 0.51 |

[a] Effect estimates (coefficient) are difference in mean (BMI SD units) or odds ratio per SD increase in genetic risk score; [b] for the binary outcomes pseudo-$R^2$ are presented; [c] for BMI F-statistic is presented; for binary outcomes AUC is presented. Education = high education vs low education around the time of pregnancy; Parity = multiparous vs nulliparous; Diabetes = Any diabetes vs none; Smoking = Any smoking during pregnancy yes vs no; Alcohol = Any alcohol consumption during pregnancy yes vs no. Abbreviations: BMI, body mass index; GRS, genetic risk score; CI, confidence interval; ALSPAC, Avon Longitudinal Study of Parents and Children; BiB, Born in Bradford; MoBa. Norwegian Mother, Father and Child Cohort Study.

**Table S4.** Exploring associations between the smoking GRS (lifetime smoking index) and risk factors for CHDs. We also include the association of the smoking GRS with smoking (also shown in table X within the manuscript) for comparison.

| Risk factor | N | Coefficient (95% CI)[a] | P-value | $R^2$ / Pseudo $R^2$ [b] | F statistic [c] | AUC |
|---|---|---|---|---|---|---|
| **ALSPAC** | | | | | | |
| Smoking | 6,428 | 1.27 (1.20, 1.35) | $1 \times 10^{-16}$ | 1.6% | - | 0.56 |
| Education | 6,806 | 0.84 (0.79, 0.90) | $3 \times 10^{-7}$ | 0.68% | - | 0.55 |
| Parity | 6,982 | 1.01 (0.96, 1.05) | 0.81 | 0.001% | - | 0.50 |
| Diabetes | 6,786 | 0.92 (0.66, 1.28) | 0.61 | 0.06% | - | 0.54 |
| Alcohol | 6,087 | 1.00 (0.94, 1.06) | 0.92 | 0.0002% | - | 0.50 |
| BMI | 6,253 | 0.06 (0.03, 0.08) | $3 \times 10^{-6}$ | 0.35% | 22 | - |
| **BiB** | | | | | | |
| Smoking | 6,482 | 1.36 (1.27, 1.45) | $2 \times 10^{-20}$ | 2.2% | - | 0.59 |
| Education | 6,483 | 0.95 (0.90, 1.00) | 0.04 | 0.09% | - | 0.52 |
| Parity | 7,259 | 0.96 (0.91, 1.00) | 0.06 | 0.06% | - | 0.51 |
| Diabetes | 7,133 | 0.93 (0.86, 1.01) | 0.10 | 0.08% | - | 0.52 |
| Alcohol | 2,110 | 0.98 (0.90, 1.07) | 0.67 | 0.01% | - | 0.50 |
| BMI | 6,196 | 0.03 (0.001, 0.05) | 0.04 | 0.07% | 4 | - |
| **MoBa** | | | | | | |
| Smoking | 20,981 | 1.23 (1.17, 1.29) | $7 \times 10^{-17}$ | 0.8% | - | 0.56 |
| Education | 21,921 | 0.87 (0.85, 0.90) | $9 \times 10^{-22}$ | 0.6% | - | 0.54 |
| Parity | 23,869 | 1.01 (0.98, 1.04) | 0.48 | 0.003% | - | 0.50 |
| Diabetes | 23,869 | 1.02 (0.91, 1.13) | 0.75 | 0.003% | - | 0.51 |
| Alcohol | 19,737 | 0.99 (0.96, 1.02) | 0.48 | 0.004% | - | 0.50 |
| BMI | 22,533 | 0.04 (0.03, 0.06) | $1 \times 10^{-10}$ | 0.2% | 42 | - |

[a] Effect estimates (coefficient) are difference in mean (BMI SD units) or odds ratio per SD increase in genetic risk score; [b] for the binary outcomes pseudo-$R^2$ are presented; [c] for BMI F-statistic is presented; for binary outcomes AUC is presented. Education = high education vs low education around the time of pregnancy; Parity = multiparous vs nulliparous; Diabetes = Any diabetes vs none; Smoking = Any smoking during pregnancy yes vs no; Alcohol = Any alcohol consumption during pregnancy yes vs no. Abbreviations: BMI, body mass index; GRS, genetic risk score; CI, confidence interval; ALSPAC, Avon Longitudinal Study of Parents and Children; BiB, Born in Bradford; MoBa. Norwegian Mother, Father and Child Cohort Study.

**Table S5.** Exploring associations between the alcohol GRS (drinks per week) and risk factors for CHDs. We also include the association of the alcohol GRS with alcohol (also shown in table X within the manuscript) for comparison.

| Risk factor | N | Coefficient (95% CI)[a] | P-value | $R^2$ / Pseudo $R^2$ [b] | F statistic [c] | AUC |
|---|---|---|---|---|---|---|
| **ALSPAC** | | | | | | |
| Alcohol | 6,087 | 1.14 (1.07, 1.21) | $3 \times 10^{-5}$ | 0.4% | - | 0.53 |
| Education | 6,806 | 1.05 (0.98, 1.12) | 0.19 | 0.05% | - | 0.51 |
| Parity | 6,982 | 1.02 (0.98, 1.07) | 0.32 | 0.02% | - | 0.51 |
| Diabetes | 6,786 | 1.22 (0.86, 1.73) | 0.26 | 0.31% | - | 0.57 |
| Smoking | 6,428 | 1.08 (1.02, 1.14) | 0.01 | 0.15% | - | 0.52 |
| BMI | 6,253 | 0.001 (-0.02, 0.03) | 0.92 | 0.0002% | 0.01 | - |
| **BiB** | | | | | | |
| Alcohol | 2,110 | 1.08 (0.99, 1.18) | 0.09 | 0.2% | - | 0.52 |
| Education | 6,483 | 0.98 (0.93, 1.03) | 0.34 | 0.02% | - | 0.51 |
| Parity | 7,259 | 1.05 (1.00, 1.10) | 0.03 | 0.09% | - | 0.52 |
| Diabetes | 7,133 | 1.07 (0.99, 1.17) | 0.09 | 0.09% | - | 0.52 |
| Smoking | 6,482 | 0.84 (0.79, 0.89) | $4 \times 10^{-8}$ | 0.8% | - | 0.55 |
| BMI | 6,196 | -0.02 (-0.05, 0.01) | 0.16 | 0.03% | 2 | - |
| **MoBa** | | | | | | |
| Alcohol | 19,737 | 1.02 (0.99, 1.05) | 0.13 | 0.02% | - | 0.51 |
| Alcohol sensitivity [d] | 19,737 | 1.06 (1.01, 1.10) | 0.01 | 0.07% | - | 0.52 |
| Education | 21,921 | 1.00 (0.96, 1.02) | 0.58 | 0.002% | - | 0.50 |
| Parity | 23,869 | 1.00 (0.98, 1.03) | 0.93 | <0.0001% | - | 0.50 |
| Diabetes | 23,869 | 1.00 (0.89, 1.10) | 0.88 | 0.0007% | - | 0.50 |
| Smoking | 20,981 | 1.02 (0.97, 1.07) | 0.37 | 0.009% | - | 0.51 |
| BMI | 22,533 | -0.007 (-0.019, 0.006) | 0.32 | 0.004% | 1 | - |

[a] Effect estimates (coefficient) are difference in mean (BMI SD units) or odds ratio per SD increase in genetic risk score; [b] for the binary outcomes pseudo-$R^2$ are presented; [c] for BMI F-statistic is presented; for binary outcomes AUC is presented. Education = high education vs low education around the time of pregnancy; Parity = multiparous vs nulliparous; Diabetes = Any diabetes vs none; Smoking = Any smoking during pregnancy yes vs no; Alcohol = Any alcohol consumption during pregnancy yes vs no. Abbreviations: BMI, body mass index; GRS, genetic risk score; CI, confidence interval; ALSPAC, Avon Longitudinal Study of Parents and Children; BiB, Born in Bradford; MoBa. Norwegian Mother, Father and Child Cohort Study.

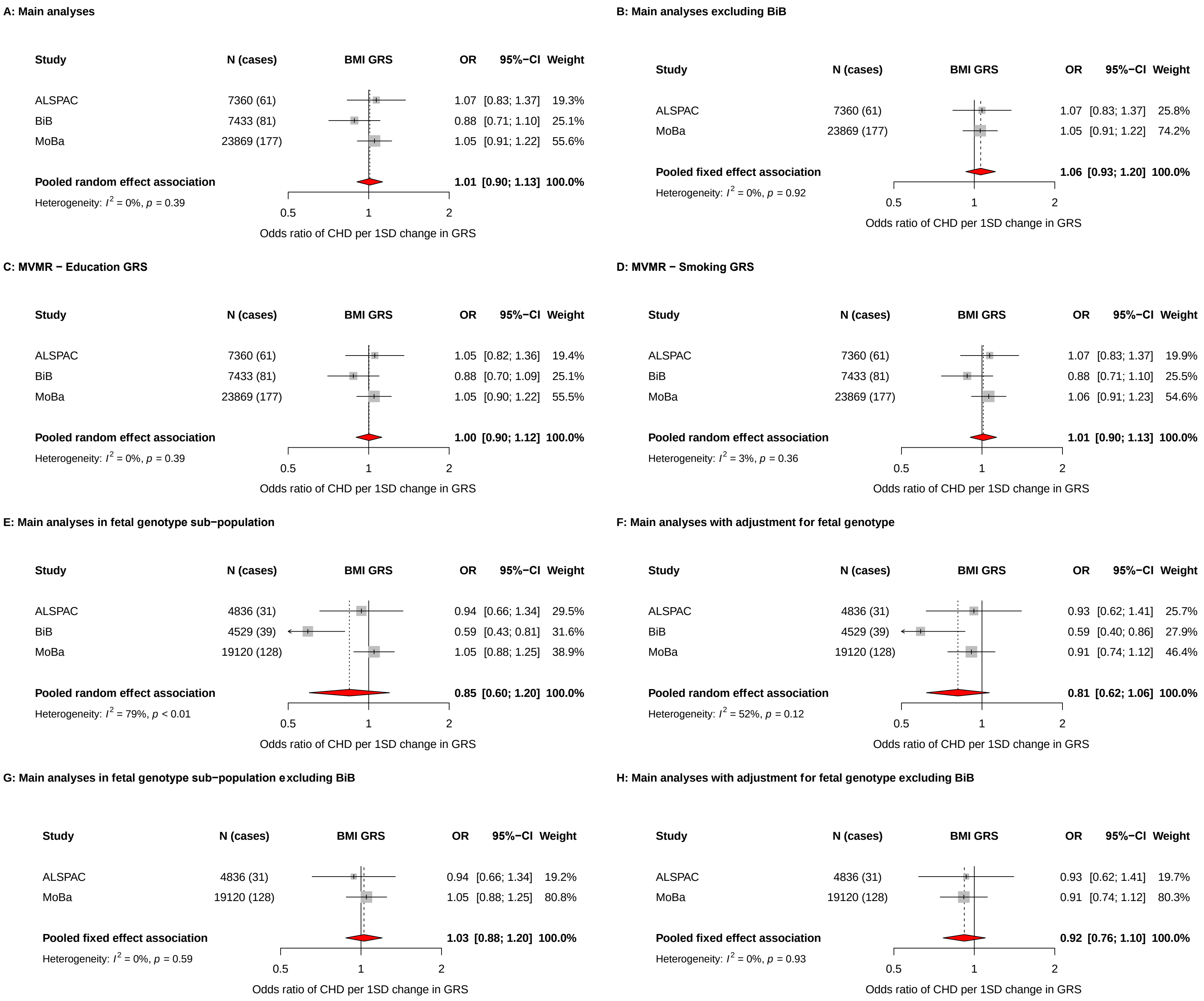
A: Main analyses
Study N (cases) BMI GRS OR 95%−CI Weight
ALSPAC 7360 (61) 1.07 [0.83; 1.37] 19.3%
BiB 7433 (81) 0.88 [0.71; 1.10] 25.1%
MoBa 23869 (177) 1.05 [0.91; 1.22] 55.6%
Pooled random effect association 1.01 [0.90; 1.13] 100.0%
Heterogeneity: $I^2$ = 0%, $p$ = 0.39
0.5 1 2
Odds ratio of CHD per 1SD change in GRS
B: Main analyses excluding BiB
Study N (cases) BMI GRS OR 95%−CI Weight
ALSPAC 7360 (61) 1.07 [0.83; 1.37] 25.8%
MoBa 23869 (177) 1.05 [0.91; 1.22] 74.2%
Pooled fixed effect association 1.06 [0.93; 1.20] 100.0%
Heterogeneity: $I^2$ = 0%, $p$ = 0.92
0.5 1 2
Odds ratio of CHD per 1SD change in GRS
C: MVMR − Education GRS
Study N (cases) BMI GRS OR 95%−CI Weight
ALSPAC 7360 (61) 1.05 [0.82; 1.36] 19.4%
BiB 7433 (81) 0.88 [0.70; 1.09] 25.1%
MoBa 23869 (177) 1.05 [0.90; 1.22] 55.5%
Pooled random effect association 1.00 [0.90; 1.12] 100.0%
Heterogeneity: $I^2$ = 0%, $p$ = 0.39
0.5 1 2
Odds ratio of CHD per 1SD change in GRS
D: MVMR − Smoking GRS
Study N (cases) BMI GRS OR 95%−CI Weight
ALSPAC 7360 (61) 1.07 [0.83; 1.37] 19.9%
BiB 7433 (81) 0.88 [0.71; 1.10] 25.5%
MoBa 23869 (177) 1.06 [0.91; 1.23] 54.6%
Pooled random effect association 1.01 [0.90; 1.13] 100.0%
Heterogeneity: $I^2$ = 3%, $p$ = 0.36
0.5 1 2
Odds ratio of CHD per 1SD change in GRS
E: Main analyses in fetal genotype sub−population
Study N (cases) BMI GRS OR 95%−CI Weight
ALSPAC 4836 (31) 0.94 [0.66; 1.34] 29.5%
BiB 4529 (39) 0.59 [0.43; 0.81] 31.6%
MoBa 19120 (128) 1.05 [0.88; 1.25] 38.9%
Pooled random effect association 0.85 [0.60; 1.20] 100.0%
Heterogeneity: $I^2$ = 79%, $p$ < 0.01
0.5 1 2
Odds ratio of CHD per 1SD change in GRS
F: Main analyses with adjustment for fetal genotype
Study N (cases) BMI GRS OR 95%−CI Weight
ALSPAC 4836 (31) 0.93 [0.62; 1.41] 25.7%
BiB 4529 (39) 0.59 [0.40; 0.86] 27.9%
MoBa 19120 (128) 0.91 [0.74; 1.12] 46.4%
Pooled random effect association 0.81 [0.62; 1.06] 100.0%
Heterogeneity: $I^2$ = 52%, $p$ = 0.12
0.5 1 2
Odds ratio of CHD per 1SD change in GRS
G: Main analyses in fetal genotype sub−population excluding BiB
Study N (cases) BMI GRS OR 95%−CI Weight
ALSPAC 4836 (31) 0.94 [0.66; 1.34] 19.2%
MoBa 19120 (128) 1.05 [0.88; 1.25] 80.8%
Pooled fixed effect association 1.03 [0.88; 1.20] 100.0%
Heterogeneity: $I^2$ = 0%, $p$ = 0.59
0.5 1 2
Odds ratio of CHD per 1SD change in GRS
H: Main analyses with adjustment for fetal genotype excluding BiB
Study N (cases) BMI GRS OR 95%−CI Weight
ALSPAC 4836 (31) 0.93 [0.62; 1.41] 19.7%
MoBa 19120 (128) 0.91 [0.74; 1.12] 80.3%
Pooled fixed effect association 0.92 [0.76; 1.10] 100.0%
Heterogeneity: $I^2$ = 0%, $p$ = 0.93
0.5 1 2
Odds ratio of CHD per 1SD change in GRS

**Figure S1.** Showing the main results and results from additional analyses for the MR analyses of genetically predicted maternal BMI and offspring CHDs. Odds ratios (ORs) of CHD for a 1SD difference in maternal GRS in each study and pooled across studies using random effects meta-analysis or fixed-effects analyses when excluding BiB (panels B, G, H). Adjusted for top 10 genetic principal components in all cohorts with additional adjustment for genetic chip, genetic batch, and imputation batch in MoBa. **Panel A:** Main analyses as shown in the main manuscript. **Panel B:** Main analyses excluding BiB. **Panel C:** Main analyses with additional adjustment for genetically predicted educational attainment (Multivariable Mendelian randomization analyses). **Panel D:** Main analyses with additional adjustment for genetically predicted smoking (Multivariable Mendelian randomization analyses). **Panel E:** Main analysis results in the sub-population with fetal genotype. **Panel F:** As Panel E, but with additional adjustment for fetal genotype. **Panel G:** As panel E but excluding BiB. **Panel H:** As panel F but excluding BiB. Abbreviations: ALSPAC, Avon Longitudinal Study of Parents and Children; BiB, Born in Bradford; MoBa, Norwegian Mother, Father and Child Cohort Study; BMI, body mass index; CI, confidence interval; CHD, congenital heart disease; SD, standard deviation; GRS, genetic risk score; MVMR, multivariable Mendelian randomization.

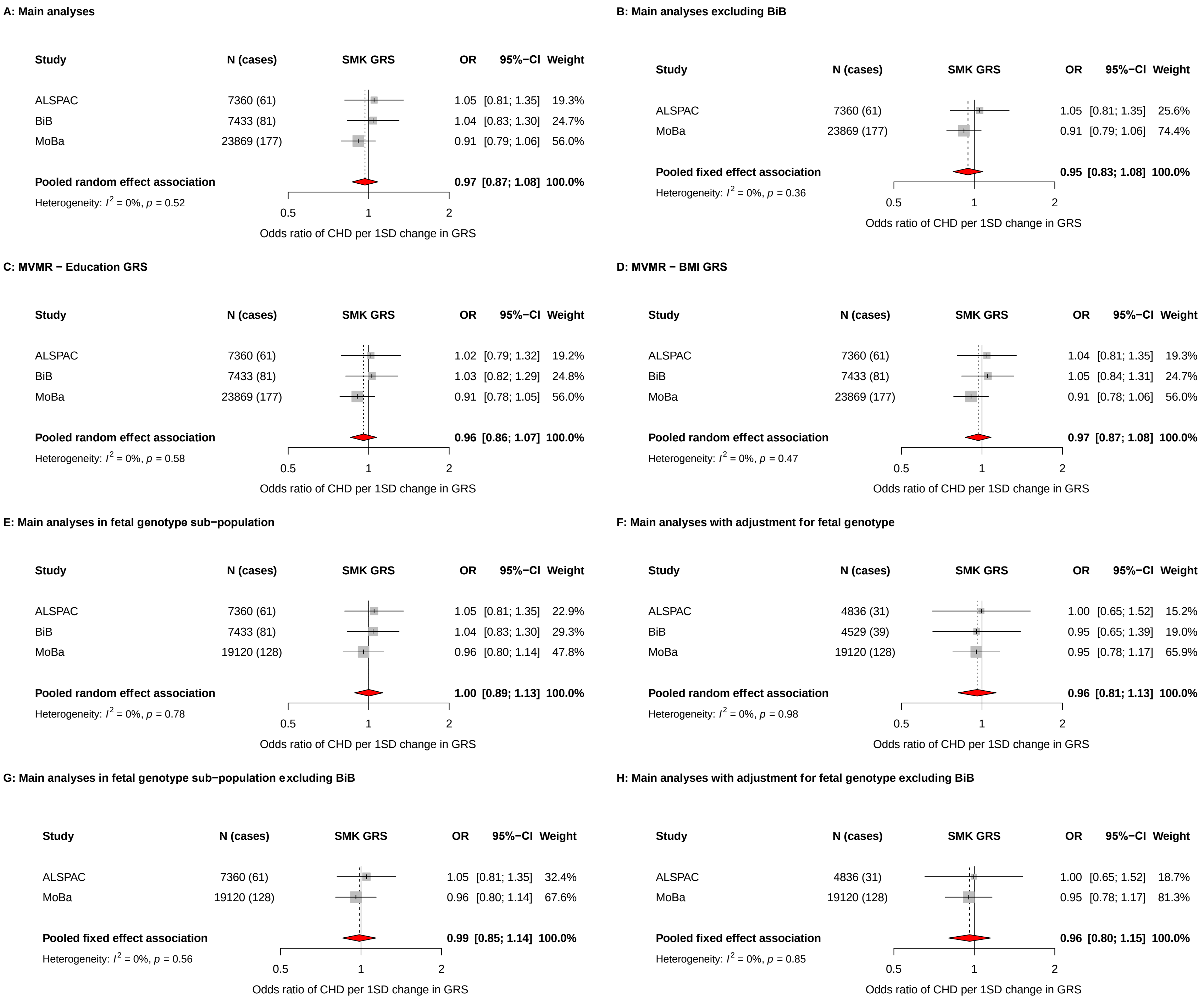
A: Main analyses
Study N (cases) SMK GRS OR 95%−CI Weight
ALSPAC 7360 (61) 1.05 [0.81; 1.35] 19.3%
BiB 7433 (81) 1.04 [0.83; 1.30] 24.7%
MoBa 23869 (177) 0.91 [0.79; 1.06] 56.0%
Pooled random effect association 0.97 [0.87; 1.08] 100.0%
Heterogeneity: $I^2$ = 0%, $p$ = 0.52
0.5 1 2
Odds ratio of CHD per 1SD change in GRS
B: Main analyses excluding BiB
Study N (cases) SMK GRS OR 95%−CI Weight
ALSPAC 7360 (61) 1.05 [0.81; 1.35] 25.6%
MoBa 23869 (177) 0.91 [0.79; 1.06] 74.4%
Pooled fixed effect association 0.95 [0.83; 1.08] 100.0%
Heterogeneity: $I^2$ = 0%, $p$ = 0.36
0.5 1 2
Odds ratio of CHD per 1SD change in GRS
C: MVMR − Education GRS
Study N (cases) SMK GRS OR 95%−CI Weight
ALSPAC 7360 (61) 1.02 [0.79; 1.32] 19.2%
BiB 7433 (81) 1.03 [0.82; 1.29] 24.8%
MoBa 23869 (177) 0.91 [0.78; 1.05] 56.0%
Pooled random effect association 0.96 [0.86; 1.07] 100.0%
Heterogeneity: $I^2$ = 0%, $p$ = 0.58
0.5 1 2
Odds ratio of CHD per 1SD change in GRS
D: MVMR − BMI GRS
Study N (cases) SMK GRS OR 95%−CI Weight
ALSPAC 7360 (61) 1.04 [0.81; 1.35] 19.3%
BiB 7433 (81) 1.05 [0.84; 1.31] 24.7%
MoBa 23869 (177) 0.91 [0.78; 1.06] 56.0%
Pooled random effect association 0.97 [0.87; 1.08] 100.0%
Heterogeneity: $I^2$ = 0%, $p$ = 0.47
0.5 1 2
Odds ratio of CHD per 1SD change in GRS
E: Main analyses in fetal genotype sub−population
Study N (cases) SMK GRS OR 95%−CI Weight
ALSPAC 7360 (61) 1.05 [0.81; 1.35] 22.9%
BiB 7433 (81) 1.04 [0.83; 1.30] 29.3%
MoBa 19120 (128) 0.96 [0.80; 1.14] 47.8%
Pooled random effect association 1.00 [0.89; 1.13] 100.0%
Heterogeneity: $I^2$ = 0%, $p$ = 0.78
0.5 1 2
Odds ratio of CHD per 1SD change in GRS
F: Main analyses with adjustment for fetal genotype
Study N (cases) SMK GRS OR 95%−CI Weight
ALSPAC 4836 (31) 1.00 [0.65; 1.52] 15.2%
BiB 4529 (39) 0.95 [0.65; 1.39] 19.0%
MoBa 19120 (128) 0.95 [0.78; 1.17] 65.9%
Pooled random effect association 0.96 [0.81; 1.13] 100.0%
Heterogeneity: $I^2$ = 0%, $p$ = 0.98
0.5 1 2
Odds ratio of CHD per 1SD change in GRS
G: Main analyses in fetal genotype sub−population excluding BiB
Study N (cases) SMK GRS OR 95%−CI Weight
ALSPAC 7360 (61) 1.05 [0.81; 1.35] 32.4%
MoBa 19120 (128) 0.96 [0.80; 1.14] 67.6%
Pooled fixed effect association 0.99 [0.85; 1.14] 100.0%
Heterogeneity: $I^2$ = 0%, $p$ = 0.56
0.5 1 2
Odds ratio of CHD per 1SD change in GRS
H: Main analyses with adjustment for fetal genotype excluding BiB
Study N (cases) SMK GRS OR 95%−CI Weight
ALSPAC 4836 (31) 1.00 [0.65; 1.52] 18.7%
MoBa 19120 (128) 0.95 [0.78; 1.17] 81.3%
Pooled fixed effect association 0.96 [0.80; 1.15] 100.0%
Heterogeneity: $I^2$ = 0%, $p$ = 0.85
0.5 1 2
Odds ratio of CHD per 1SD change in GRS

**Figure S2.** Showing the main results and results from additional analyses for the MR analyses of genetically predicted maternal smoking (using a genetic risk score of a lifetime smoking index) and offspring CHDs. Odds ratios (ORs) of CHD for a 1SD difference in maternal GRS in each study and pooled across studies using random effects meta-analysis or fixed-effects analyses when excluding BiB (panels B, G, H). Adjusted for top 10 genetic principal components in all cohorts with additional adjustment for genetic chip, genetic batch, and imputation batch in MoBa. **Panel A:** Main analyses as shown in the main manuscript. **Panel B:** Main analyses excluding BiB. **Panel C:** Main analyses with additional adjustment for genetically predicted educational attainment (Multivariable Mendelian randomization analyses). **Panel D:** Main analyses with additional adjustment for genetically predicted body mass index (Multivariable Mendelian randomization analyses). **Panel E:** Main analysis results in the sub-population with fetal genotype. **Panel F:** As Panel E, but with additional adjustment for fetal genotype. **Panel G:** As panel E but excluding BiB. **Panel H:** As panel F but excluding BiB. Abbreviations: ALSPAC, Avon Longitudinal Study of Parents and Children; BiB, Born in Bradford; MoBa, Norwegian Mother, Father and Child Cohort Study; BMI, body mass index; CI, confidence interval; CHD, congenital heart disease; SD, standard deviation; GRS, genetic risk score; MVMR, multivariable Mendelian randomization.

A: Main analyses

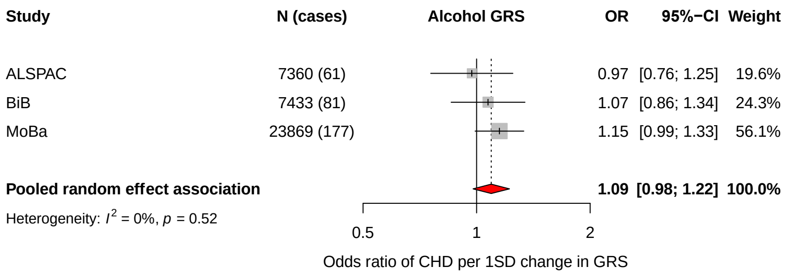


B: Main analyses excluding BiB

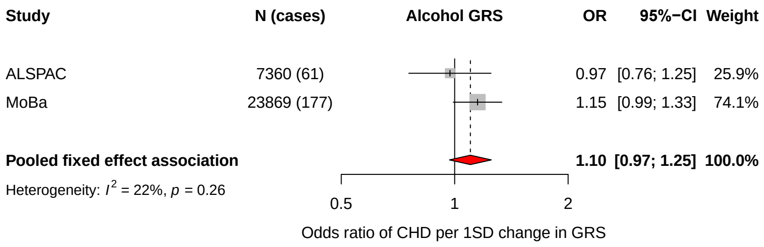


C: MVMR − Smoking GRS

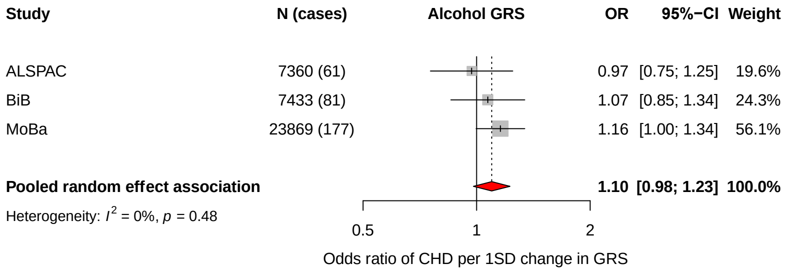


D: Main analyses in fetal genotype sub−population

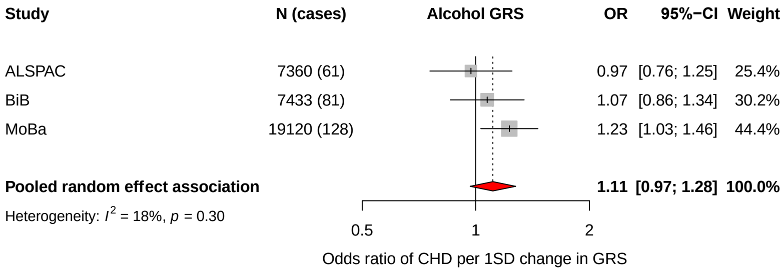


E: Main analyses with adjustment for fetal genotype

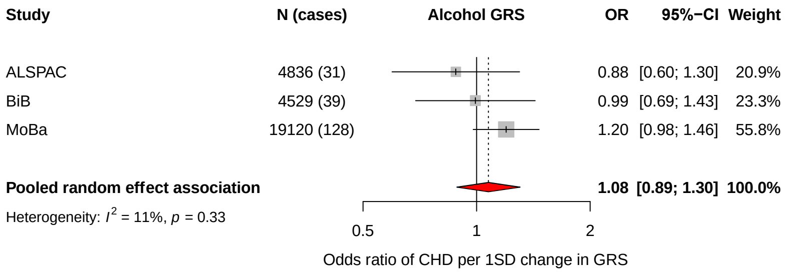


F: Main analyses in fetal genotype sub−population excluding BiB

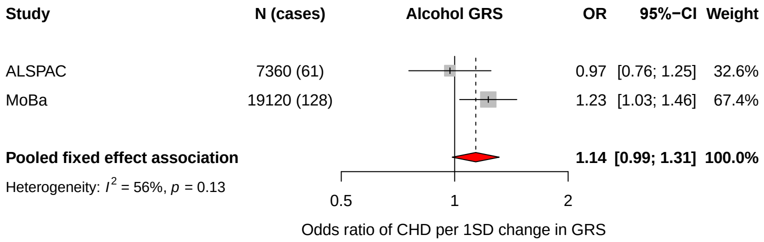


G: Main analyses with adjustment for fetal genotype excluding BiB

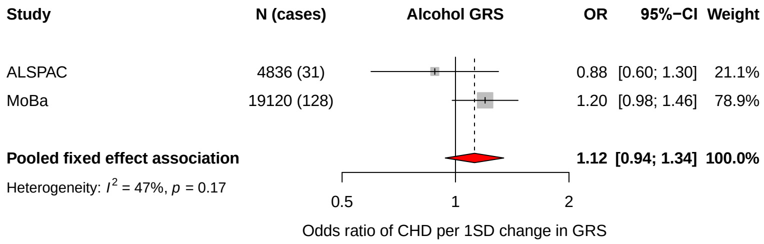

**Figure S3.** Showing the main results and results from additional analyses for the MR analyses of genetically predicted maternal alcohol consumption (using a genetic risk score of drinks per week) and offspring CHDs. Odds ratios (ORs) of CHD for a 1SD difference in maternal GRS in each study and pooled across studies using random effects meta-analysis or fixed-effects analyses when excluding BiB (panels B, G, H). Adjusted for top 10 genetic principal components in all cohorts with additional adjustment for genetic chip, genetic batch, and imputation batch in MoBa. **Panel A:** Main analyses as shown in the main manuscript. **Panel B:** Main analyses excluding BiB. **Panel C:** Main analyses with additional adjustment for genetically predicted smoking (Multivariable Mendelian randomization analyses). **Panel D:** Main analysis results in the sub-population with fetal genotype. **Panel E:** As Panel D, but with additional adjustment for fetal genotype. **Panel F:** As panel D but excluding BiB. **Panel G:** As panel E but excluding BiB. Abbreviations: ALSPAC, Avon Longitudinal Study of Parents and Children; BiB, Born in Bradford; MoBa, Norwegian Mother, Father and Child Cohort Study; BMI, body mass index; CI, confidence interval; CHD, congenital heart disease; SD, standard deviation; GRS, genetic risk score; MVMR, multivariable Mendelian randomization.

**References cited in Supplementary Material**


1. Hemani G, Zheng J, Elsworth B, Wade KH, Haberland V, Baird D, et al. The MR-Base platform supports systematic causal inference across the human phenome. Elife. 2018 May 30;7:e34408.

2. Myers TA, Chanock SJ, Machiela MJ. LDlinkR: An R Package for Rapidly Calculating Linkage Disequilibrium Statistics in Diverse Populations. Front Genet. 2020;11:157.

3. Tyrrell J, Richmond RC, Palmer TM, Feenstra B, Rangarajan J, Metrustry S, et al. Genetic Evidence for Causal Relationships Between Maternal Obesity-Related Traits and Birth Weight. JAMA [Internet]. 2016 Mar 15 [cited 2020 Nov 17];315(11):1129. Available from: http://jama.jamanetwork.com/article.aspx?doi=10.1001/jama.2016.1975

4. Corbin LJ, Richmond RC, Wade KH, Burgess S, Bowden J, Smith GD, et al. BMI as a Modifiable Risk Factor for Type 2 Diabetes: Refining and Understanding Causal Estimates Using Mendelian Randomization. Diabetes. 2016 Oct;65(10):3002–7.

5. Xu L, Borges MC, Hemani G, Lawlor DA. The role of glycaemic and lipid risk factors in mediating the effect of BMI on coronary heart disease: a two-step, two-sample Mendelian randomisation study. Diabetologia. 2017 Nov;60(11):2210–20.

6. Carter AR, Gill D, Davies NM, Taylor AE, Tillmann T, Vaucher J, et al. Understanding the consequences of education inequality on cardiovascular disease: mendelian randomisation study. BMJ. 2019 May 22;365:l1855.

7. Taylor AE, Richmond RC, Palviainen T, Loukola A, Wootton RE, Kaprio J, et al. The effect of body mass index on smoking behaviour and nicotine metabolism: a Mendelian randomization study. Hum Mol Genet. 2019 Apr 15;28(8):1322–30.

8. Carreras-Torres R, Johansson M, Haycock PC, Relton CL, Davey Smith G, Brennan P, et al. Role of obesity in smoking behaviour: Mendelian randomisation study in UK Biobank. BMJ [Internet]. 2018 May 16 [cited 2021 Dec 17];k1767. Available from: https://www.bmj.com/lookup/doi/10.1136/bmj.k1767

9. Khouja JN, Sanderson E, Wootton RE, Taylor AE, Munafò MR. A multivariable Mendelian randomisation study exploring the direct effects of nicotine on health compared with the other constituents of tobacco smoke: Implications for e-cigarette use. medRxiv [Internet]. 2021 Jan 1;2021.01.12.21249493. Available from: http://medrxiv.org/content/early/2021/01/15/2021.01.12.21249493.abstract

10. Lee JJ, Wedow R, Okbay A, Kong E, Maghzian O, Zacher M, et al. Gene discovery and polygenic prediction from a genome-wide association study of educational attainment in 1.1 million individuals. Nat Genet. 2018 Jul 23;50(8):1112–21.
